# Phase-locked transcranial electrical brain stimulation for tremor suppression in dystonic tremor syndromes

**DOI:** 10.1101/2021.11.03.21265869

**Authors:** Freek Nieuwhof, Ivan Toni, Arthur W.G. Buijink, Anne-Fleur van Rootselaar, Bart P.C. van de Warrenburg, Rick C. Helmich

**Affiliations:** Centre for Cognitive Neuroimaging, Donders Institute for Brain, Cognition and Behaviour, Radboud University, Nijmegen, The Netherlands; Department of Neurology, Center of Expertise for Parkinson & Movement Disorders, Donders Institute for Brain, Cognition and Behaviour, Radboud University Medical Centre, Nijmegen, The Netherlands; Department of Neurology and Clinical Neurophysiology, Amsterdam University Medical Centers, Amsterdam Neuroscience, University of Amsterdam, Amsterdam, the Netherlands

**Keywords:** Tremor, Dystonia, electrical stimulation, transcranial alternating current stimulation

## Abstract

**Background:** Tremor is a common and burdensome symptom in patients with dystonia, which is clinically heterogeneous and often resistant to treatment. The pathophysiology is suggested to involve abnormal activity in the cerebellum and motor cortex, but the causal role of these brain regions remains to be established. Transcranial alternating current stimulaton (TACS) can suppress rhytmic cerebral activity in other tremor disorders when phase-locked to the ongoing arm tremor, but the effect on dystonic tremor syndromes is unknown.

**Objective/Hypothesis:** We aimed to establish the causal role of the cerebellum and motor cortex in dystonic tremor syndromes, and explore the therapeutic efficacy of phase-locked TACS.

**Methods:** We applied phase-locked TACS over the ipsilateral cerebellum (N=14) and contralateral motor cortex (N=17) in dystonic tremor syndrome patients, while patients assumed a tremor-evoking posture. We measured tremor power using accelerometery during 30s stimulation periods at 10 different phase-lags (36-degrees increments) between tremor and TACS for each target. Post-hoc, TACS-effects were related to a key clinical feature: the jerkiness (regularity) of tremor.

**Results:** Cerebellar TACS modulated tremor amplitude in a phase-dependent manner, such that tremor amplitude was suppressed or enhanced at opposite sides of the phase-cycle. This effect was specific for patients with non-jerky (sinusoidal) tremor (n=10), but absent in patients with jerky (irregular) tremor (n=4). Phase-locked stimulation over the motor cortex did not modulate tremor amplitude.

**Conclusions:** This study indicates that the cerebellum plays a causal role in the generation of (non-jerky) dystonic tremor syndrome. Our findings suggest pathophysiologic heterogeneity between patients with dystonic tremor syndrome, which mirrors clinical variability.

## Introduction

Tremor, defined as an involuntary, rhythmic, oscillatory movement of a body part [1] is a highly burdensome symptom present in 17-55% of patients with dystonia [2-4]. In dystonia, tremor can be present in the body part affected by dystonia, defined as dystonic tremor, or in a non-dystonic body part, defined as “tremor associated with dystonia” [1]. Clinically, dystonic tremor syndromes typically occur during actions, although sometimes also at rest. Furthermore, the tremor can be posture-dependent or task-specific, and it may be jerky (irregular) in nature [5]. Some have argued that jerky “tremor” in dystonia is a misnomer, since the (irregular) nature does not fit the classical definition of (regular) tremor [6].

Current treatments, such as botulinum toxin, pharmacological interventions or deep brain stimulation (DBS), are invasive and/or based on trial-and-error, often with unsatisfactory results [7,8]. These unsatisfactory effects are likely due to inadequate targeting of the underlying tremor circuit. It has been hypothesized that dystonic tremor syndromes involve abnormal activity in the cerebello-thalamo-cortical circuit and basal ganglia [9-11]. Involvement of the cerebello-thalamo-cortical circuit is supported by several lines of research. Stereotactic interventions in the ventral intermediate nucleus of the thalamus (VIM), a cerebellar relay nucleus, led to tremor suppression in dystonic tremor syndromes [8,12-15]. Furthermore, functional MRI during a grip-force task has been used as a proxy of tremor-related cerebral activity [16], showing similar pathological grip-force related activity in the cerebellum between dystonic and essential tremor. Since cerebellar dysfunction is well-established in essential tremor [17-19], this suggests that cerebellar dysfunction may also play a role in dystonic tremor syndromes. Finally, classical eye blink conditioning, a marker of cerebellar dysfunction, is abnormal in patients with dystonic tremor syndromes [20]. These findings suggest that the cerebello-thalamo-cortical circuit is involved in dystonic tremor syndromes, with possible additional involvement of other circuits depending on clinical variability within this group (i.e. the basal ganglia in jerky tremor) [10,21]. Causal evidence for the involvement of the cerebello-thalamo-cortical circuit in dystonic tremor syndromes is lacking. Here, we aimed to reduce tremor amplitude by suppressing tremulous activity in two nodes of this circuit, i.e. the motor cortex and cerebellum, using transcranial alternating current stimulation (TACS).

TACS enables non-invasive modulation of brain oscillations in a frequency and phase-specific manner [22]. By applying electrical currents to the brain, TACS can rhythmically modulate membrane potentials, bringing it further from or closer to the firing threshold [23-25]. TACS can be personalized by using the frequency and/or phase of the tremor, measured with accelerometery, as a proxy for pathological cerebral oscillations. In tremor types other than dystonic tremor syndromes, TACS has been used to entrain tremor phase [26]. For example, TACS at individual tremor frequencies entrained essential tremor when applied over the cerebellum [27], and it entrained physiological tremor when applied over the (pre-)motor cortex [28,29] or cerebellum [29]. Furthermore, TACS reduced tremor amplitude when phase-locked to the ongoing tremor. This has been shown in Parkinson’s tremor with stimulation of the motor cortex [30] and in essential tremor with stimulation of the cerebellum [31]. In Parkinson’s tremor, specific phase-lags between ongoing tremor and TACS led to reduced tremor amplitude, while opposite phase-lags led to augmented tremor amplitude [30]. In contrast, in essential tremor, phase-locked TACS reduced tremor compared to sham, but effects were not specific to a certain phase-lag [31]. Furthermore, the effect of TACS in essential tremor depended on tremor phenotype, with TACS being more effective for patients with a regular, stable tremor [31].

Given the involvement of the cerebello-thalamo-cortical circuit in dystonic tremor syndromes, we tested the hypothesis that phase-locked TACS over the cerebellum and motor cortex attenuates tremor amplitude. Based on recent findings in other tremor disorders, TACS-related tremor attenuation might be phase-specific (as observed in Parkinson’s tremor [30]) or phase-independent (as observed in essential tremor [31]). Finally, we explored whether the effects of TACS depend on inter-individual differences in tremor phenotype. We focused on the jerkiness of the tremor, because: (i) jerkiness is a well-known clinical feature of some dystonic tremor syndromes [5]; (ii) the presence/absence of jerkiness of dystonic tremor is associated with different neural responses in the globus pallidus [21], and (iii) our intervention (TACS, which is per definition sinusoidal) may be more effective to a non-jerky (regular) versus a jerky (irregular) tremor [31].

## Material and methods

### Participants

We included 17 participants with a dystonic tremor syndrome from the Radboud University Medical center (Radboudumc 12 participants) and the Amsterdam UMC, location AMC (5 participants). Inclusion criteria were a clinical diagnosis of dystonic tremor or tremor associated with dystonia, with the presence of primary focal or segmental dystonia, according to the most recent consensus statement [1]. Patients with questionable dystonia were classified as essential tremor plus and excluded [1,32]. Further exclusion criteria were: a history of traumatic brain injury or stroke, moderate to severe head tremor when lying supine, signs of myoclonus dystonia, and the use of anti-tremor medication other than propranolol. Propranolol was tapered off for the patients using it (N=5) in the week prior to measurements, so all measurements were done in the OFF-medication state. To prevent any effect of botulinum toxin treatment, measurements for a single participant who received injections in the most tremulous arm were scheduled six months after the latest injection. All participants participated voluntarily and gave their written informed consent prior to starting the study. The included 17 patients resulted from 30 initial referrals. Two experienced movement disorder neurologists (RH and BvdW) carefully examined video recordings of these 30 patients. This led to two patients being excluded because of diagnostic doubt (suspicion of functional tremor and myoclonus dystonia), two because of a high perceived burden of participation and nine because tremor was very subtle and/or inconsistently present. The study was approved by the ethical committee ‘Commissie mensgebonden onderzoek (CMO) regio Arnhem-Nijmegen’ and was performed according to the principles of the Declaration of Helsinki.

### Experimental procedures and design

Participants were first clinically examined. Table 1 provides details on the clinical testing. Then, we asked participants to lie down supine on a treatment table and placed a tri-axial accelerometer (Brain Products) on their most tremulous hand. We placed the accelerometer on the location where tremor was best captured (dorsum of the hand or one of the fingers). We determined the posture of the most affected arm in which tremor was most pronounced, and patients were instructed to assume this specific posture during the remainder of the experiment. The least-affected arm was always in rest.

**Table 1.** Participant characteristics.

| Participant number | Motor cortex stimulation | Cerebellum stimulation | Age (years) | Gender (M/F) | Age tremor onset | BFM | Dystonia in most tremulous arm | Cervical dystonia | TRS total | TRS most affected arm | Tremor frequency (Hz) | Rest tremor | Head tremor | Jerky tremor | Asymmetric tremor | Postural dependency | FAB | MOCA |
| --- | --- | --- | --- | --- | --- | --- | --- | --- | --- | --- | --- | --- | --- | --- | --- | --- | --- | --- |
| 1 | yes | no | 60.0 | F | 30.0 | 27.5 | yes | yes | 40 | 13 | 4.8 | yes | yes | yes | no | yes | 11 | 23 |
| 2 | yes | yes | 70.0 | F | 31.0 | 4.0 | yes | yes | 22 | 10 | 5.6 | yes | yes | no | yes | no | 16 | 27 |
| 3 | yes | yes | 64.0 | F | 51.0 | 8.0 | yes | yes | 58 | 22 | 5.0 | yes | yes | no | yes | yes | 18 | 26 |
| 4 | yes | yes | 62.0 | F | 45.0 | 9.0 | yes | yes | 54 | 22 | 4.2 | yes | yes | yes | no | yes | 18 | 30 |
| 5 | yes | yes | 58.0 | M | 45.0 | 4.0 | yes | yes | 39 | 15 | 5.8 | yes | no | no | yes | no | 17 | 25 |
| 6 | yes | yes | 69.0 | M | 60.0 | 16.0 | yes | yes | 39 | 18 | 4.2 | yes | no | yes | yes | no | 15 | 26 |
| 7 | yes | yes | 74.0 | F | 40.0 | 1.5 | yes | yes | 39 | 13 | 4.6 | yes | yes | no | yes | yes | 17 | 28 |
| 8 | yes | yes | 74.0 | M | 55.0 | 0.5 | no | yes | 43 | 16 | 5.6 | yes | no | no | yes | yes | 18 | 30 |
| 9 | yes | yes | 67.0 | F | 55.0 | 3.0 | yes | yes | 35 | 19 | 5.4 | yes | no | no | yes | yes | 14 | 23 |
| 10 | yes | no | 78.0 | M | 20.0 | 1.0 | yes | no | 45 | 19 | 5.0 | yes | no | yes | yes | no | 17 | 28 |
| 11 | yes | yes | 82.0 | M | 70.0 | 8.5 | yes | yes | 55 | 20 | 4.0 | yes | no | no | yes | yes | 13 | 19 |
| 12 | yes | no | 28.0 | F | 11.0 | 13.5 | yes | yes | 36 | 9 | 9.4 | yes | no | yes | yes | no | - | - |
| 13 | yes | yes | 56.0 | M | 48.0 | 8.5 | yes | yes | 34 | 18 | 5.0 | yes | no | no | yes | no | 17 | 31 |
| 14 | yes | yes | 75.0 | M | - | 7.0 | no | yes | 84 | 23 | 4.6 | yes | yes | no | yes | no | 17 | 25 |
| 15 | yes | yes | 60.0 | M | 52.0 | 7.0 | yes | no | 18 | 11 | 6.4 | no | no | no | yes | yes | 17 | 29 |
| 16 | yes | yes | 37.0 | M | 16.0 | 5.0 | yes | yes | 37 | 15 | 7.0 | yes | no | yes | no | yes | 12 | 28 |
| 17 | yes | yes | 68.0 | F | 20.0 | 2.0 | yes | no | 31 | 18 | 5.0 | yes | no | yes | yes | yes | 17 | 30 |
| Median (range) or count | 17 yes/0 no | 14 yes/3 no | 67.0 (28-82) | 9M/8 F | 45 (11-70) | 7.0 (0.5-27.5) | 15 yes/2 no | 14yes /3no | 39 (18-84) | 18 (9-23) | 5.0 (4.0-9.4) | 16 yes/1 no | 6 yes /11 no | 7 yes /10 no | 13yes /3 no | 10yes /7 no | 17.0 (11-18) | 27.0 (19-31) |

During the main experiment, we applied phase-locked TACS over two targets: motor cortex contralateral to the most-affected arm and cerebellum ipsilateral to the most-affected arm (order randomized between participants). For both targets, TACS was applied during 10 trials with set phase lags between ongoing tremor and stimulation (36-degree increments, randomized order). For each trial, participants assumed the predetermined tremor-evoking posture with their most affected hand. When tremor was present, TACS started with a 5s ramp up period, followed by 30 seconds of phase-locked stimulation and a 5s ramp down period (Fig 1a). After each trial participants rested their arm until they were ready for the next trial. Motor cortex stimulation was done for all participants, while cerebellum stimulation was done for 14 participants (3 missing due to time limitations). Participants were blinded to the set phase-lag.

**Figure 1.**
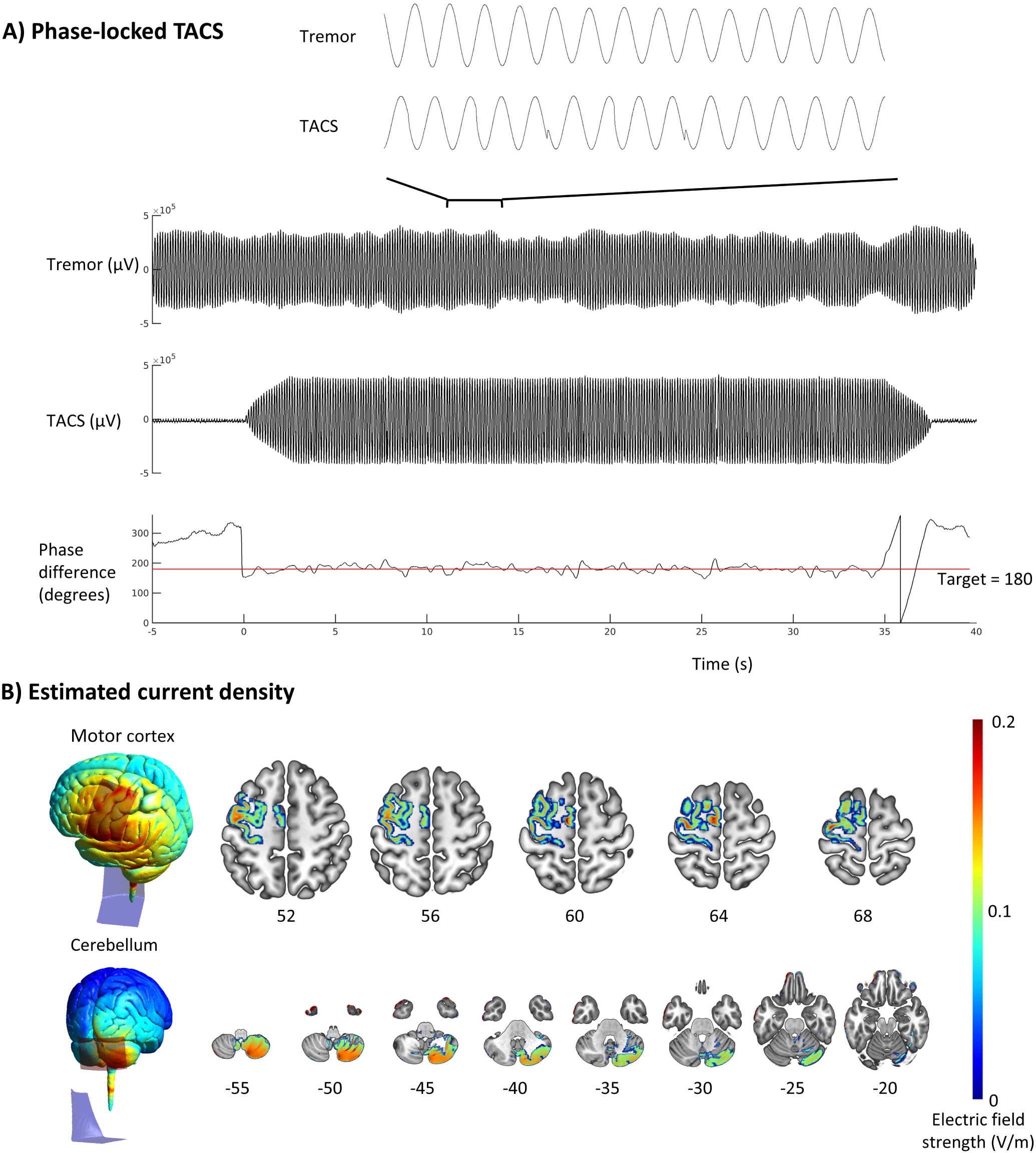
Phase-locked transcranial alternating current stimulation. Transcranial alternating current stimulation (TACS) was applied over motor cortex and cerebellum while participants were holding their most affected hand in a tremor evoking posture. (A) Prior to each trial, participants lifted their hand in the predefined posture. When tremor was present, TACS was applied at a set phase lag from ongoing tremor (180 degrees in this example), with 5s ramp up, 30s of stimulation and 5s of ramp down. The shown “tremor” signal is the accelerometery signal of the axis with highest tremor power. (B) Simulations of the electric field (V/m) in the Simnibs MNI template for motor cortex and cerebellum montages. Electric fields in axial slices are thresholded at 0.1 V/m and masked by grey matter in Brodmann areas 4 and 6 of the Brainnetome atlas [58] for the motor cortex montage or grey matter for the cerebellum montage. Numbers under axial slices represent MNI z-coordinates.

### Phase-locked transcranial alternating current stimulation

For motor cortex stimulation, one electrode (5×7 cm) was centred on the contralateral (to tremor) C3/C4 location as localized using the EEG 10/20 system, with a second electrode (5×11 cm) on the ipsilateral (to tremor) shoulder [30,33]. For cerebellum stimulation, one electrode was centred 3 cm lateral of the inion (ipsilateral to tremor), with a second electrode (5×11 cm) located on the contralateral (to tremor) shoulder [34,35]. Electrodes were attached using Ten20 paste (MedCat) and impedance was kept below 10 kOhm (motor cortex 5.3 ± 2.9 kOhm, Cerebellum 5.1 ± 2.6 kOhm). Current densities (Fig. 1b) were simulated using SIMNIBS version 3.2 [36], with the template head model and standard conductivities provided by SIMNIBS.

A custom-built system continuously traced frequency and phase of the accelerometery axis with highest tremor amplitude (as determined during posture selection) and generated a stimulation signal with equal frequency and set phase lag from the accelerometery signal (supplementary figure 1). This stimulation signal was fed into a DC-stimulator PLUS (Eldith, NeuroConn GmbH, Germany), which then provided phase-locked TACS with a peak-to-peak stimulation current of 2 mA (Fig. 1a). We used a BrainAmp ExG amplifier (Brain Products GmbH, Gilching, Germany) for simultaneous recording of accelerometery and TACS-output signals (sampling frequency 5kHz).

### Data processing

Data was processed offline in MATLAB (The Mathworks Inc., version R2018a, Natrick, RI, USA) using Fieldtrip [37]. Accelerometery data of the axis on which phase-locked stimulation was based (i.e. the axis with highest tremor power) was first bandpass filtered between 2 and 20 Hz. We then segmented the data into 5s segments over which we calculated an averaged power spectrum with a 0.2 Hz resolution to determine peak tremor frequency. We then further bandpass filtered the accelerometery data with a 2Hz passband centred around the peak tremor frequency [30]. Next, we applied Hilbert transformation to obtain the amplitude and phase of tremor (i.e. the filtered accelerometery signal) and the phase of the applied TACS-signal [30]. Tremor amplitude (the absolute value of the Hilbert transformation) was z-scored relative to the mean and standard deviation of all ramp-up periods for each target (motor cortex or cerebellum) [31]. We averaged the z-scored tremor amplitude over the full 30s stimulation period for the analysis on phase-lag specific modulation of tremor amplitude and over the first and second half (15s) of stimulation for the analysis on phase-independent modulation of tremor amplitude.

To quantify the degree of phase-locking, the phase-locking value (PLV) was calculated between accelerometery and TACS. We calculated the PLV over the initial 10s of phase-locked stimulation only (ramp-up and first 5s of stable stimulation), since successful tremor suppression would result in subtle or even absent tremor that is untraceable.

### Statistical analysis

For each trial, we statistically tested whether we achieved the set phase-lag between tremor and TACS with a V-test [38]. This is a test for non-uniformity of the phase-lag with a known mean direction (i.e. the set phase-lag). For one trial (subject 6, cerebellum stimulation, phase-lag 288) adequate phase-locking was not achieved (V-test p-value > 0.05). This trial was interpolated based on the two adjacent phase-lags.

To test whether we achieved phase-lag specific modulation of tremor amplitude, we used a statistical approach that adequately deals with individual variation in preferred phase, as was observed in phase-locked TACS in Parkinson’s tremor [30], and is highly sensitive to detect phase-dependent effects [39]. We fitted two models to the individual phase-lag vs. tremor amplitude (average z-score over 30s stimulation periods) response curves: one intercept only model and one with intercept and two circular predictors (sine and cosine). In the case of phase-specific modulation of tremor amplitude, the model with circular predictors will better fit the data than the model without, which we tested on an individual basis with F-tests. Individual p-values resulting from this F-test were then taken to the group level with Fisher’s method [39]. We calculated effect sizes (F-squared) of the model comparison for each individual and averaged this over the group [40].

To test whether phase-locked stimulation reduced tremor amplitude independent of the phase-lag [31], we averaged the first and second halves of stimulation over all trials for each target. For each of the two data segments, we used a one-sample t-test (two-sided) to test whether tremor amplitude deviated from zero (baseline). We calculated Cohen’s d effect size for the second half of stimulation [40]. In addition, we tested whether tremor amplitude during the second data segment differed from the first data segment using a two-sample t-test. For all classical statistical tests, a p-value < 0.05 was used as threshold for statistical significance.

We supplemented classical statistical testing with Bayesian statistics using JASP Version 0.14.1 (JASP Team (2020). Bayesian statistics were added to quantify evidence in favour of either the alternative or null hypothesis. Bayes factors were interpreted according to the guidelines provided in JASP [41], in which Bayes factors (BF_10_) of 1-3, 3-10 or >10 are considered anecdotal, moderate or strong evidence for the alternative hypothesis, while BF_10_ of 0.33-1, 0.1-0.33 or <0.1 are considered anecdotal, moderate or strong evidence for the null hypothesis.

In addition, we tested whether stimulation effects (percentage change in tremor amplitude at the optimal phase difference and individual effect sizes) were correlated with achieved phase locking accuracy (PLV), dystonia severity (BFM score, most-affected hand), tremor severity (TRS score, most-affected hand), using Spearman’s Rho correlation coefficients. Furthermore, we post-hoc explored whether the stimulation effects depended on the jerkiness of tremor (clinical consensus by RH and BvdW). Previous studies in essential tremor showed more reliable phase-tracking [42] and more amplitude modulation by TACS [31] in patients with more regular (non-jerky) tremor. We focused this analysis on cerebellum stimulation (n=14), given the large effect size and anecdotal Bayesian evidence in favour of a phase-dependent effect on tremor amplitude. There were four patients with a jerky tremor and 10 patients with a non-jerky tremor. We validated the clinical observation of jerkiness by comparing the frequency variability (frequency width half magnitude, FWHM) [43] between the two groups, using a Mann-Whitney U test. Furthermore, we compared individual effect sizes (F-squared) between jerky and non-jerky tremor using a Mann-Whitney U test and repeated the phase-lag specific analyses including only the non-jerky group. Finally, we compared jerky and non-jerky tremor patients on dystonia severity, (BFM), tremor severity (TRS score, most-affected hand), tremor frequency, years since tremor onset and age, using Mann-Whitney U tests.

## Results

### Participant characteristics (Table 1)

Participants were 28 to 82 years old (9 men, 8 women). Two participants did not have dystonia in the hand most affected by tremor (labelled as tremor associated with dystonia), while all others did (labelled as dystonic tremor) [1]. Rest tremor was present in all but one participant, postural tremor was clearly asymmetric in 13 participants, and tremor was clinically judged as jerky in 7 out of 17 participants.

The Fahn-Tolosa-Marin Tremor Rating Scale (TRS) parts A, B and C were assessed for clinical tremor severity (possible ranges 0-88, 0-36 and 0-28 [44]. The sum score for items covering the most tremulous hand was calculated and used as measure for tremor severity of the most affected hand (possible range 0-28). Peak tremor frequency was assessed by using accelerometery. The Burke-Fahn-Marsden rating scale (BFM, possible range 0-120) was used for severity of dystonia [45], the Montreal Cognitive Assessment (MOCA, possible range 0-31) score for general cognitive functioning [46,47], the Frontal Assessment Battery (FAB, possible range 0-18) for frontal lobe functioning [48,49].

### Phase-locked stimulation

Phase-locked stimulation was adequately achieved in all participants (Fig. 2 and Supplementary material). Mean PLVs were 0.83 (0.55-0.94) in motor cortex stimulation and 0.81 (0.49-0.96) in cerebellum stimulation (median, range). We observed a significant correlation between mean PLV and overall tremor amplitude for motor cortex stimulation (Spearman’s Rho=0.59, p=0.01), indicating that phase-locked stimulation was more adequate with higher tremor amplitude. This correlation was not present for cerebellum stimulation (Spearman’s Rho=0.03, p=0.93).

**Figure 2.**
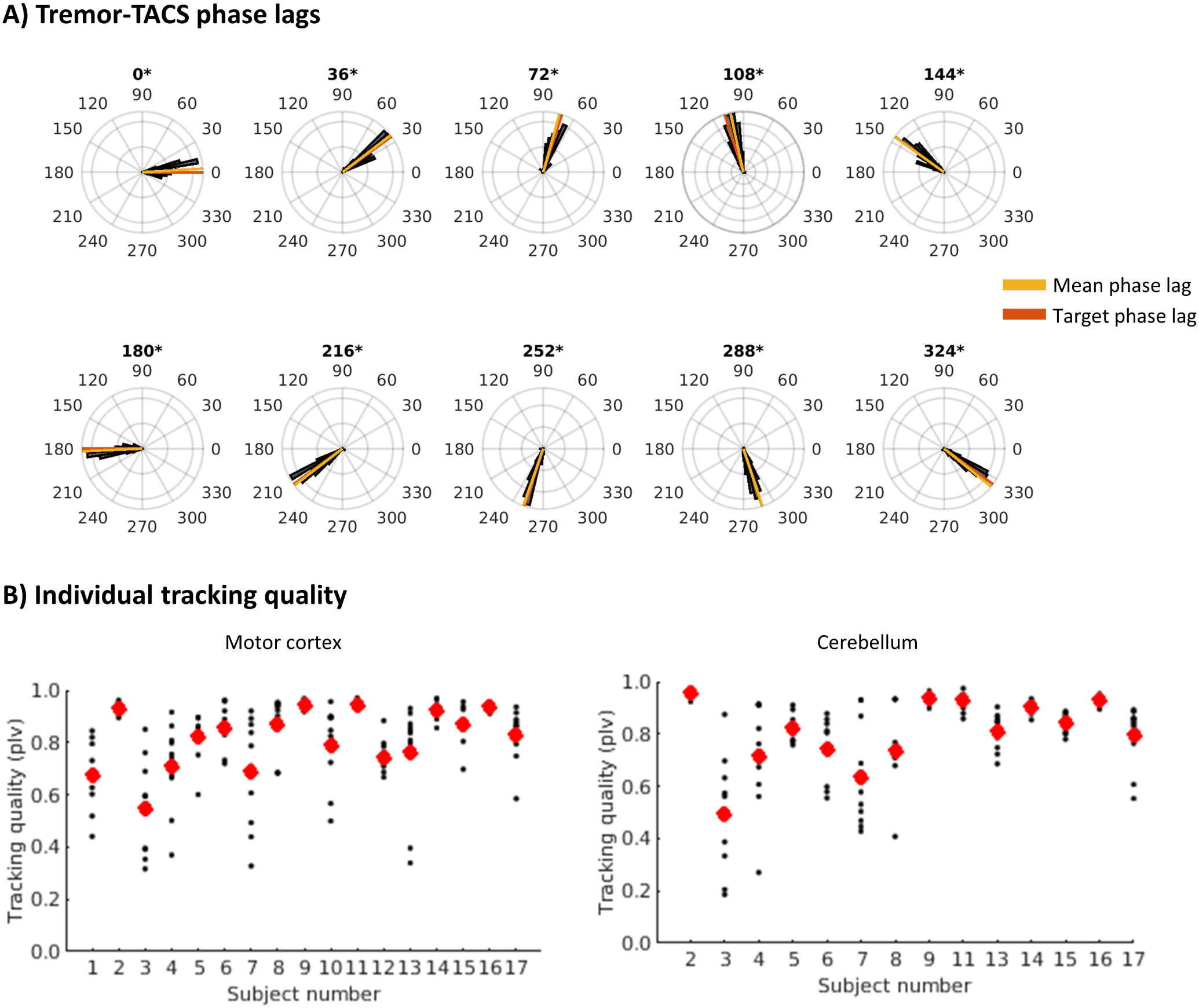
Adequacy of phase-locked stimulation. (A) Polar histograms of the phase lag between ongoing tremor and TACS for each of the ten set phase lags for a representative participant. Asterisks indicate non-uniformity with mean direction in line with the set phase lag (V-test p-value < 0.05). Polar histograms of all participants and stimulation conditions are include in the supplementary material. (B) PLVs between tremor and TACS for all participants (participant numbers match those in Table 1). Small black dots represent individual trials and large red dots represent the mean PLV over all trials. TACS = Transcranial alternating current stimulation, PLV = Phase locking value.

### Phase-dependent tremor amplitude modulation

After alignment of each patient’s data to the phase-lag with highest tremor amplitude reduction, average tremor power at that “optimal” phase-lag was, not surprisingly, significantly lower than during ramp-up (Fig. 3a, motor cortex: median change in amplitude versus ramp-up = -43.39% (range: -0.55 to -94.98%); t=-6.8, p<0.001; cerebellum: -48.48% (range: -4.47 to -96.18%); t=-5.8, p=0.001; p-values FDR corrected for 10 comparisons). Without multiple comparison correction, the phase lags of 108 degrees (t=2.5 p=0.023) for motor cortex stimulation and -144 (t=2.5, p=0.028) and 180 (t=2.4, p=0.033) degrees for cerebellum stimulation showed increased amplitude relative to ramp-up. At the group level, we observed no significant phase-specific modulation of tremor amplitude (i.e. the circular predictor model did not fit the data better than the null model) for both motor cortex and cerebellum stimulation (combined Fisher’s p=0.95 and 0.13, respectively). Effect sizes were small for motor cortex (median = 0.12; range: 0-0.92), but large for cerebellum stimulation (median = 0.42; range: 0.06-1.20). Similarly, Bayesian statistics on the group level (linear regression on median z-scores after alignment (Fig. 3a), resulted in a Bayes factor of 0.46 for motor cortex stimulation (anecdotal evidence for the null model without circular predictors) and a Bayes factor of 1.52 for cerebellum stimulation (anecdotal evidence for the model with circular predictors). For motor cortex stimulation, more accurate phase-locking (higher PLV) was related to more tremor amplitude reduction at the optimal phase-lag (Spearman’s Rho=0.49, p=0.046). No other correlations between percentage reduction or individual effect size of phase-specific modulation with dystonia severity (BFM), tremor severity (TRS), or PLV were found.

**Figure 3.**
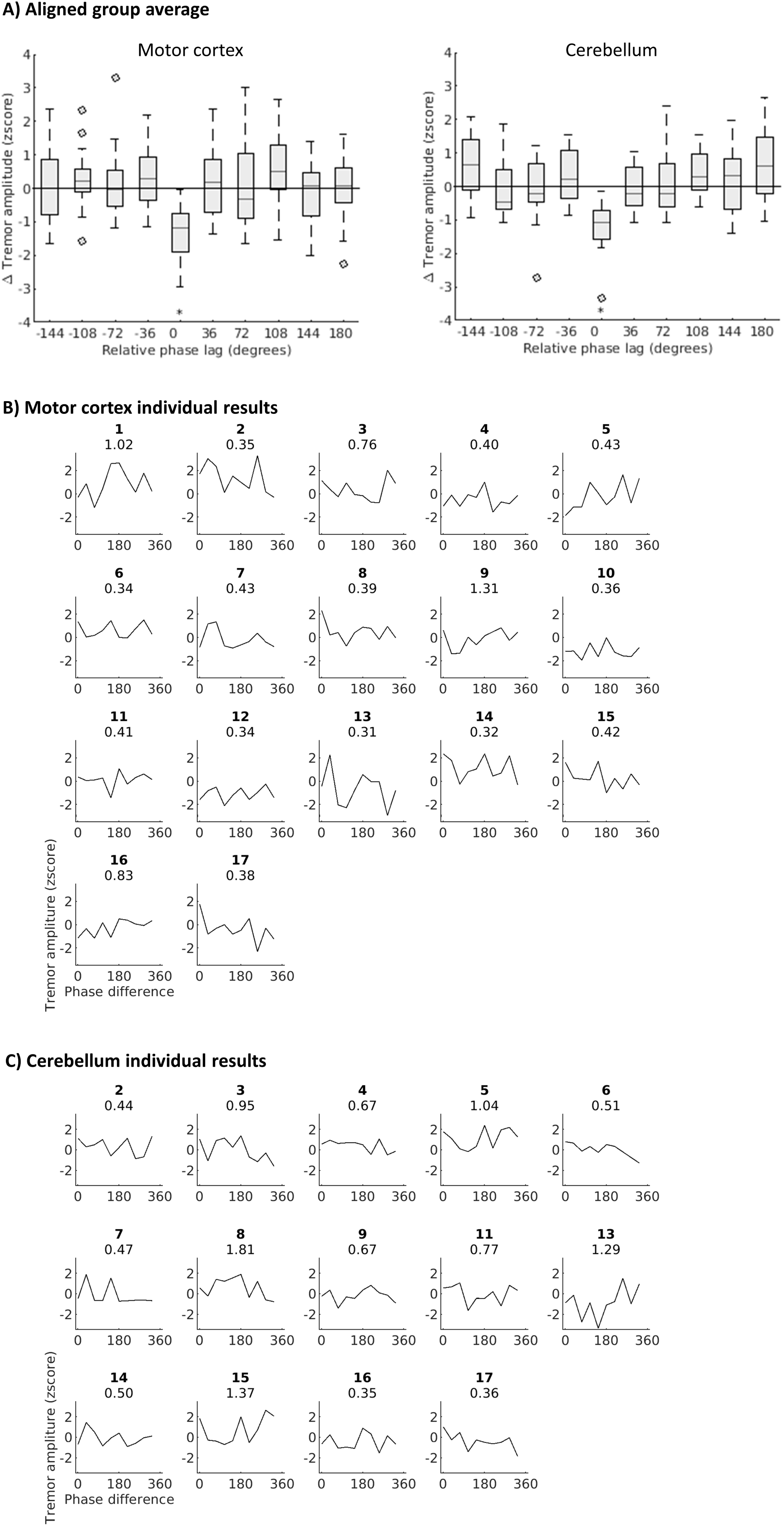
Phase-specific modulation of tremor amplitude. (A) Delta tremor amplitude (30s of stimulation – ramp up) for each phase-lag after alignment on largest tremor reduction (set to phase lag 0) for motor cortex and cerebellum stimulation. Asterisks represent significant deviation from baseline (zero). (B) Individual phase-lag vs. tremor amplitude response curves for motor cortex stimulation. Top numbers represent participant numbers (matching Table 1) and Bayes Factors in favour of the model with intercept and circular predictors vs. the null (intercept only) model. (C) as B, for cerebellum stimulation.

### Phase-dependent tremor amplitude modulation in sinusoidal (regular) tremor

Patients with jerky tremor had more frequency variability (larger FWHM) than patients with sinusoidal, regular (non-jerky) tremor (U=32, p=0.034, Fig. 4a,b). The effect sizes of phase-dependent amplitude modulation tended to be larger in patients with non-jerky (n=10) versus jerky (n=4) tremor, but this was not significant (U=6, p=0.054, Fig. 4c). Given the strong trend, we next explored the effects in the non-jerky tremor group only. After alignment to the optimal phase-lag, tremor amplitude was significantly reduced (compared to ramp-up) at this phase-lag (median change in amplitude versus ramp-up = -48.48% (range: -4.47 to -90.18%); t=-6.8 p=0.020, FDR corrected) and increased at the opposite phase differences (−144 and 180 degrees, both p=0.043, FDR corrected). In addition, we observed significant phase-dependent modulation of tremor amplitude with cerebellum stimulation in non-jerky tremor (combined Fisher’s p=0.049, Bayes factor 2.42). Dystonia severity, tremor severity, tremor frequency, years since tremor onset, and age did not differ between patients with jerky versus non-jerky tremor (all p-values >0.3).

**Figure 4.**
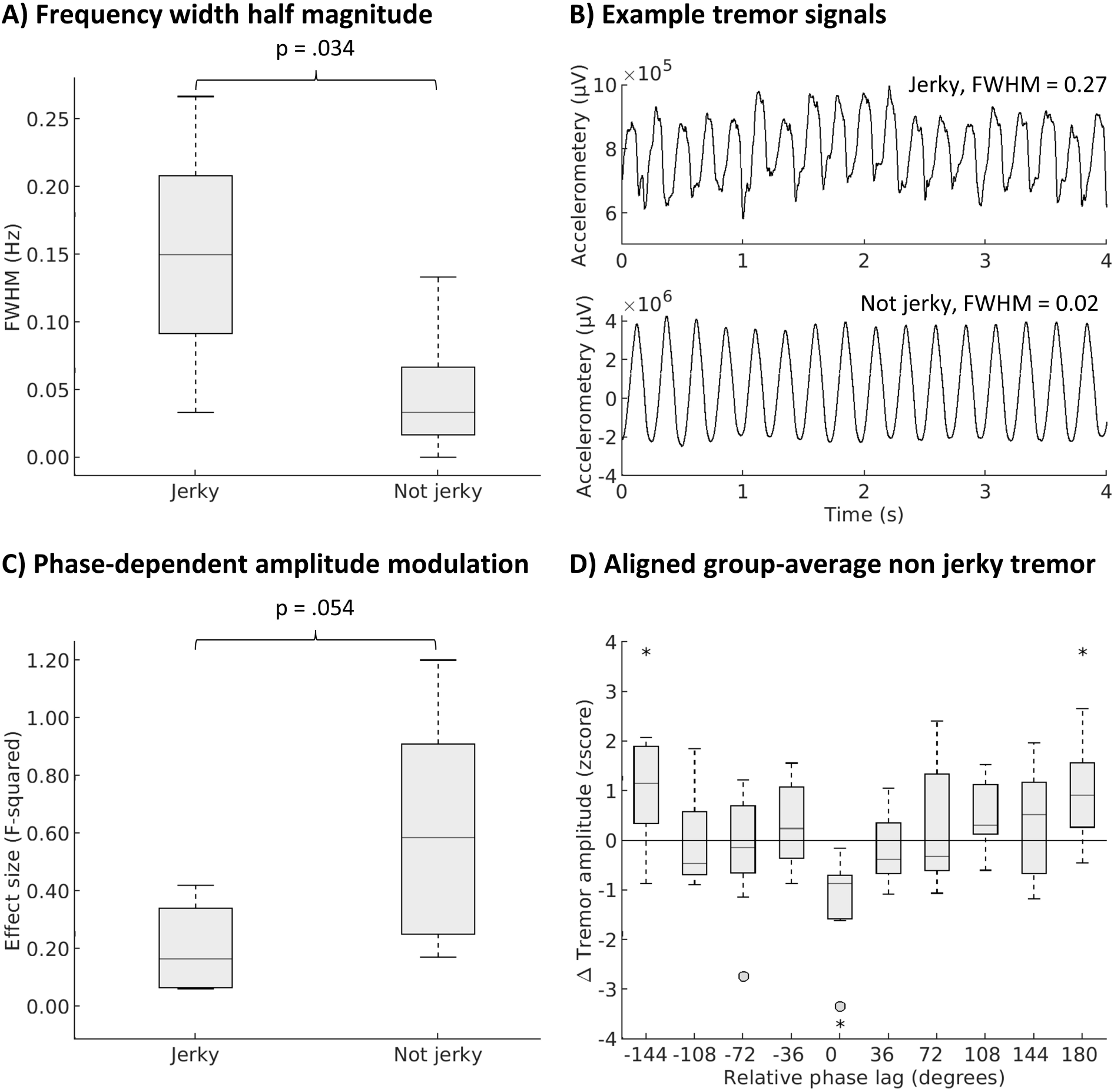
Phase-specific modulation of tremor amplitude in non-jerky tremor. (A) Frequency width half magnitude (FWHM) in jerky vs. non-jerky tremor. (B) Example tremor accelerometery signals of two patients with jerky (top) and non-jerky (bottom) tremor. (C) effect size (F-squared) of phase-dependent tremor amplitude modulation in jerky vs. non-jerky tremor. (D) Tremor amplitude aligned on phase lag with lowest value (relative phase lag 0), asterisks indicate significant deviation from zero (FDR corrected for 10 comparisons).

### Phase-independent tremor amplitude modulation

When averaging over all 10 phase-locked stimulation trials for each target, tremor amplitude was not reduced during the first or second half of the stimulation periods, with no differences between the two stimulation periods (Fig. 5, all p-values >0.05). Effect sizes for the second half of stimulation periods were small for both motor cortex (Cohen’s d = 0.003) and cerebellum stimulation (Cohen’s d = 0.132) [40]. Bayesian statistics showed moderate evidence in favour of the null hypothesis (lack of tremor reduction) for both motor cortex and cerebellum stimulation (H0: zscore=0, H1 zscore<0, Bayes factors 0.17 and 0.25 for first and second half of motor cortex stimulation periods and 0.15 and 0.20 for first and second half of cerebellum stimulation periods).

**Figure 5.**
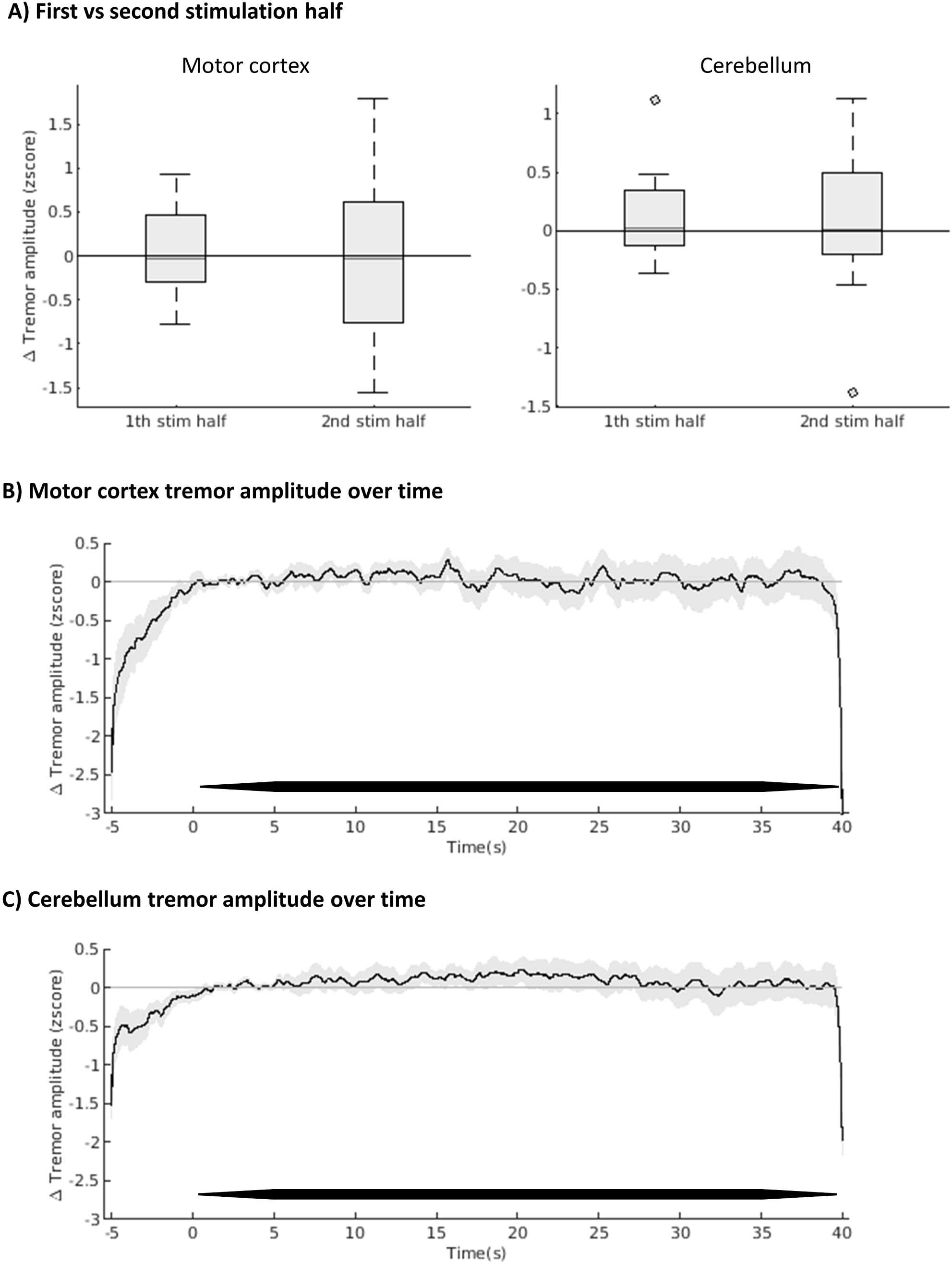
Modulation of tremor amplitude over all phase-lags. (A) Delta tremor amplitude averaged over first and second halves of stimulation periods of all trials, independent of set phase-lag. (B) Delta tremor amplitude over time, averaged over all phase-lags and participants. Black line represents mean zscore, grey lines standard error of mean. TACS is indicated by the thick black lines just above the x-axis. (C) as B, for cerebellum stimulation.

### Adverse events

The most frequently reported adverse events were pain in the neck due to lying supine (N=5) and tingling or stinging sensations underneath the stimulation electrodes (both N=5). All adverse events (Supplementary Table 1) abated directly after stopping stimulation or within hours after measurements.

## Discussion

We investigated whether phase-locked TACS over the motor cortex and cerebellum modulates tremor amplitude in dystonic tremor syndromes. Across all patients, there were no significant effects of phase-locked TACS on tremor amplitude for either region. However, in a subgroup of patients with non-jerky (regular) tremor (n=10), phase-locked TACS over the cerebellum suppressed tremor amplitude in a phase-specific manner. There were no phase-independent effects of phase-locked TACS on tremor amplitude, in contrast to findings in essential tremor [31].

These findings complement accumulating evidence that the cerebellum plays a key role in dystonic tremor syndromes [9], as it does in dystonic symptoms [50,51]. This study provides novel evidence showing that tremor amplitude can be modulated through non-invasive electrical stimulation of the cerebellum. This indicates that the cerebellum plays a causal role in dystonic tremor syndromes. Interestingly, the effect of cerebellum stimulation was specific to patients with a regular, sinusoidal (non-jerky) tremor. This suggests that clinical differences in tremor phenotype are mirrored by pathophysiologic heterogeneity. Further support for this view comes from pallidal single neuron recordings during DBS [21]. In pure dystonia and dystonia with jerky head tremor, relatively more burst cells were present, while relatively more pause cells were present in dystonia with sinusoidal head tremor. When combined with our findings, this may suggest that jerky tremor relies on pallidal alterations, while non-jerky (sinusoidal) tremor relies on cerebellar alterations. Although this is highly speculative at this stage, since there were also other differences between the previous study and this one, it may suggest that better clinical (or electrophysiologic) phenotyping of dystonic tremor syndromes may have therapeutic consequences. For example, DBS is typically most effective for dystonic tremor when applied at the border between VIM (cerebellar receiving thalamic nucleus) and VOp (pallidal receiving thalamic nucleus) [11]. Our results suggest that dystonic tremor syndrome patients with a sinusoidal tremor may benefit more from interventions in the cerebello-thalamic circuit (VIM) whereas patients with jerky tremor may benefit more from interventions in the pallido-thalamic circuit (VOp). This could be tested in future studies.

When we qualitatively compare our findings to previous comparable studies in other tremor disorders, some interesting differences can be discerned. We found no phase-dependent amplitude modulation with motor cortex stimulation, which was observed in Parkinson’s tremor [30]. This may reflect a relatively large subcortical drive in dystonic tremor syndromes compared to Parkinson’s tremor. Another qualitative discrepancy with previous studies is the lack of phase-independent tremor amplitude reduction observed here, which was present in essential tremor [31]. Although we did not directly compare them, these discrepancies suggest that there are pathophysiological differences between essential tremor, Parkinson’s tremor, and dystonic tremor syndrome. Typically, dystonic tremor is more variable and unstable when compared to essential tremor [43]. In essential tremor, non-responders to phase-locked TACS over the cerebellum had less stable tremor than responders [31]. Furthermore, in responders, amplitude reduction was associated with a reduction of temporal coherence of tremor [31]. This suggests that TACS suppressed essential tremor by disrupting stable oscillatory activity. Likewise, we observed more effect of TACS in patient with non-jerky (more stable) tremor than in patients with jerky (less stable) tremor. Phase-specific stimulation was adequate in both groups (Fig. 2B and supplementary material - patients 4,6,16 and 17 had jerky tremor), which suggests that the differential effects reflect pathophysiological rather than methodological differences. TACS may thus be most effective for patients with relatively stable tremor phenotype and stable cerebral oscillations, since those are more easily altered by TACS than more unstable cerebral oscillations.

Some methodological issues should be considered when interpreting the results of this study. First, it could be questioned whether the electrode montages we applied resulted in sufficient current densities in the targeted regions, and our set-up did not include electrophysiologic or imaging read-outs to quantify stimulation effects in the target regions. Animal studies suggest that current densities of 0.2 V/m or stronger are required for neural entrainment [52-54], with a higher chance to bias spike timing when the frequency of the external current matches the endogenous rhythm as in our case [54]. The modelling results indicate relatively low current densities (0.1-0.2 V/m, Fig. 1). However, our montages were identical to previous studies, which successfully modulated tremor in other disorders with the same stimulation intensity [27,29,30]. Second, it might be possible that we did not target the most relevant brain regions, since no study yet directly measured tremor-related brain activity in dystonic tremor syndromes. This is especially relevant for cerebellum stimulation, given that the cerebellum is positioned further away from the surface of the skull than the motor cortex. The estimated electric field with our cerebellum montage (Fig. 1) was limited to secondary somatomotor regions of the cerebellum [55]. In dystonic tremor, grip-force related activity (which is a proxy for tremor-related activity), was observed only in the primary somatomotor cerebellum [16], in line with studies in Parkinson’s tremor and essential tremor [17,56]. Hence, stimulation might be even more effective when targeting the primary somatomotor cerebellum. Future studies could optimise targeting by using higher stimulation intensities (>2 mA) to entrain more neurons [25] or use alternative head-head [31] or focused [57] electrode montages. Third, due to restricted time and burden on participants, we limited our design to a single trial for each phase-lag without a sham condition. We did not include a sham condition, because we focused on testing for phase-dependent effects [30], where all trials served as active controls to each other. Finally, it might be argued that the study is underpowered, especially the group with jerky tremor. Our findings suggest that more precise clinical phenotyping, rather than sample size per se, is an important element to consider in future studies.

## Conclusions

Phase-locked TACS over the cerebellum modulates tremor amplitude in dystonia patients with a regular, non-jerky tremor. This points towards a causal role of the cerebellum in dystonic tremor syndromes, dependent on tremor phenotype. We propose that tremor phenotype may guide optimal intervention targets.

## Data Availability

All data produced in the present study are available upon reasonable request to the authors

## Abbreviations

BFM: Burke-Fahn-Marsden rating scale
PLV: phase locking value
DBS: Deep Brain Stimulation
FAB: Frontal Assessment Battery
FWHM: Frequency Width Half Magnitude
MOCA: Montreal Cognitive Assessment
MRI: Magnetic Resonance Imaging
TACS: Transcranial alternating current stimulation
TRS: Fahn-Tolosa-Marin Tremor Rating Scale
VIM: Ventral intermediate nucleus of the thalamus
VOp: Ventralis oralis posterior nucleus of the thalamus

## Acknowledgements

We thank our participants for their time and commitment to the study. Also, we thank Uriël Plönes for his technical support in developing phase-locked TACS.

