## Supplementary for "Phase-locked transcranial electrical brain stimulation for tremor suppression in dystonic tremor syndromes"

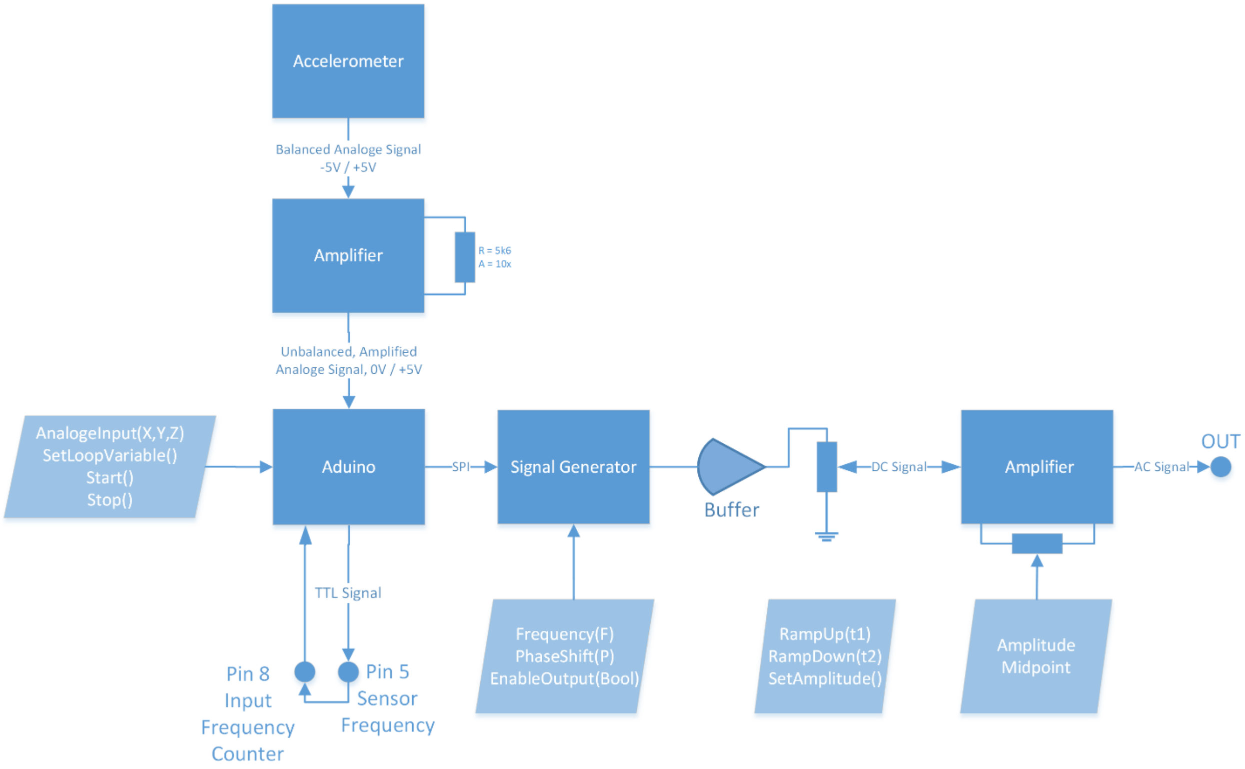


**Supplementary figure 1.**

Schematic of the custom build device used to generate phase-locked transcranial alternating current stimulation. Hardware is displayed in dark blue, settings in light blue. First, the accelerometer signal is amplified 10 times. Based on zero-crossings of this amplified signal, a block wave with equal frequency is generated. This block wave is fed into an Arduino to measure its frequency (using the FreqMeasure library function). Then, a sinusoidal signal is created in a programmable signal generator (via serial peripheral interface (SPI)), with obtained frequency and desired phase-shift. The resulting sinusoidal signal is buffered and passed though a programmable potentiometer which enables setting the ramp up, ramp down and amplitude of stimulation. The signal then passes an amplifier which gives the output signal to be fed into the DC-stimulator PLUS (Eldith, NeuroConn GmbH, Germany).

**Supplementary table 1. Numbers of participants experiencing sensations or other adverse events. All adverse events abated directly after aborting stimulation or on the same day after stopping measurements.**

| | Absent | Mild | Moderate | Severe |
|---|---|---|---|---|
| Headache | 15 | 1 | 1 | |
| Pain in neck | 12 | 5 | | |
| Pain on skull | 17 | | | |
| Tingling | 12 | 5 | | |
| Itchiness | 15 | 2 | | |
| Burning sensations | 15 | 1 | 1 | |
| Red skin under electrodes | 16 | 1 | | |
| Drowsiness | 13 | 2 | 1 | 1 |
| Loss of concentration | 15 | 2 | | |
| Suddes change of mood | 16 | 1 | | |
| Other | 11 | 6 | | |
| • Stinging sensation underneath electrode | | 5 | | |
| • Warm, pleasant feeling underneath shoulder electrode | | 1 | | |
| • Throbbing/vibrating muscle under EMG electrode | | 1 | | |

**Supplementary figures: Phase adequacy of phase-locked transcranial alternating current stimulation of the motor cortex.**

Figures on the next 9 pages show the adequacy of phase-locked stimulation for motor cortex stimulation. Figure layout is identical to figure 2 of the main manuscript.

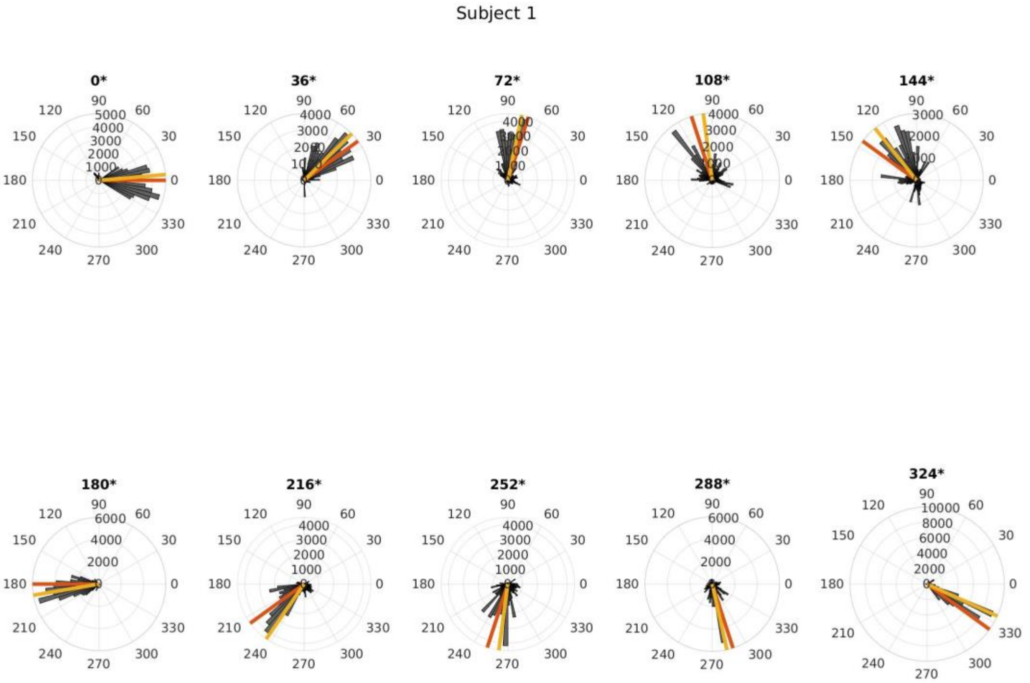
Subject 1
0*
36*
72*
108*
144*
180*
216*
252*
288*
324*

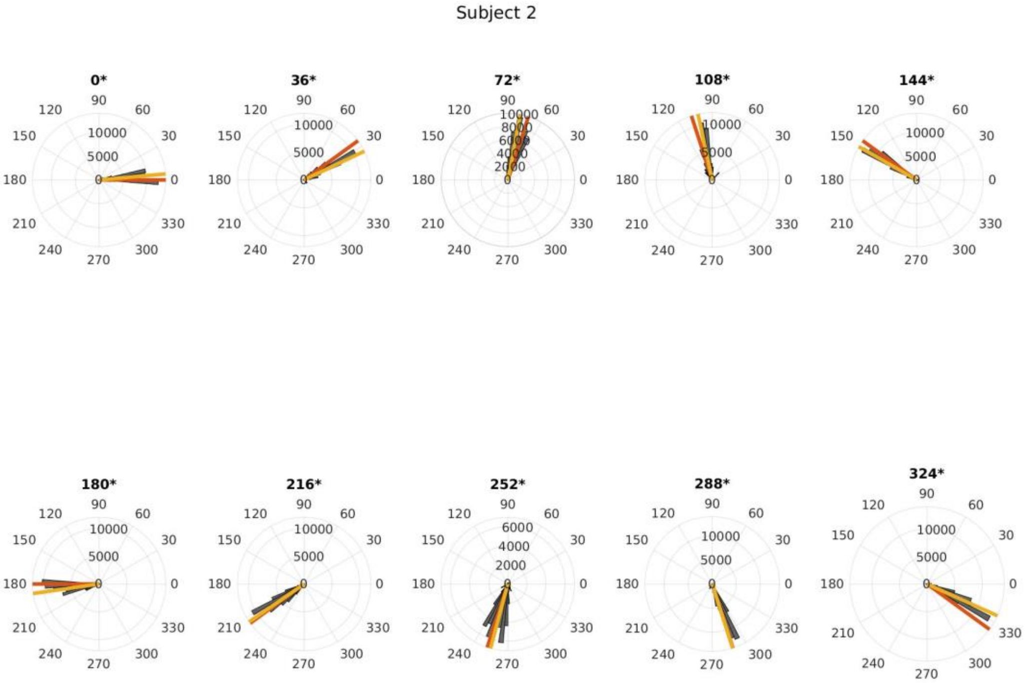
Subject 2
0*
36*
72*
108*
144*
180*
216*
252*
288*
324*

Subject 3

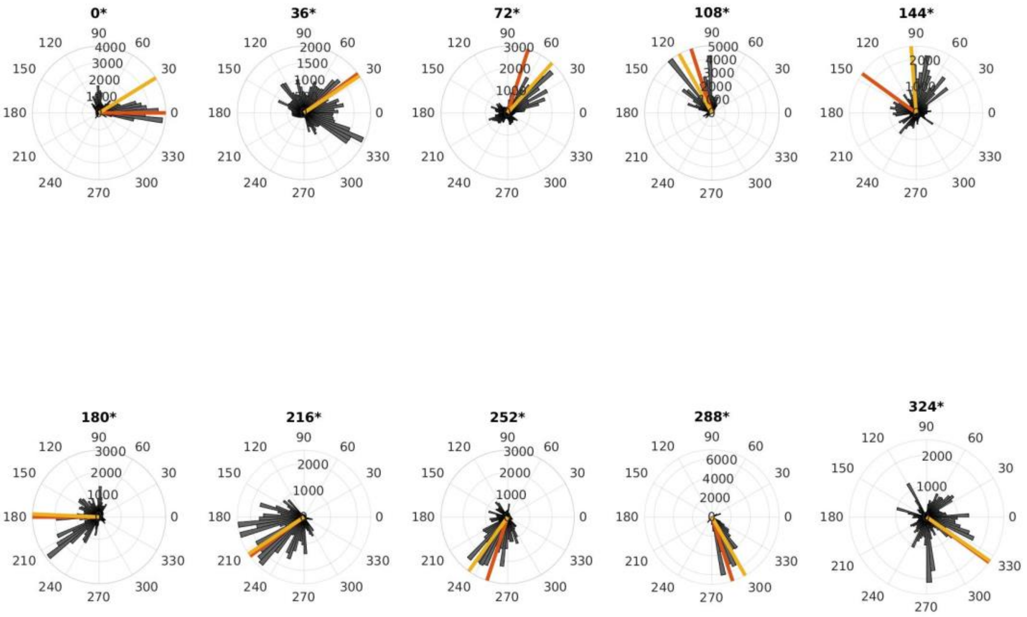


Subject 4

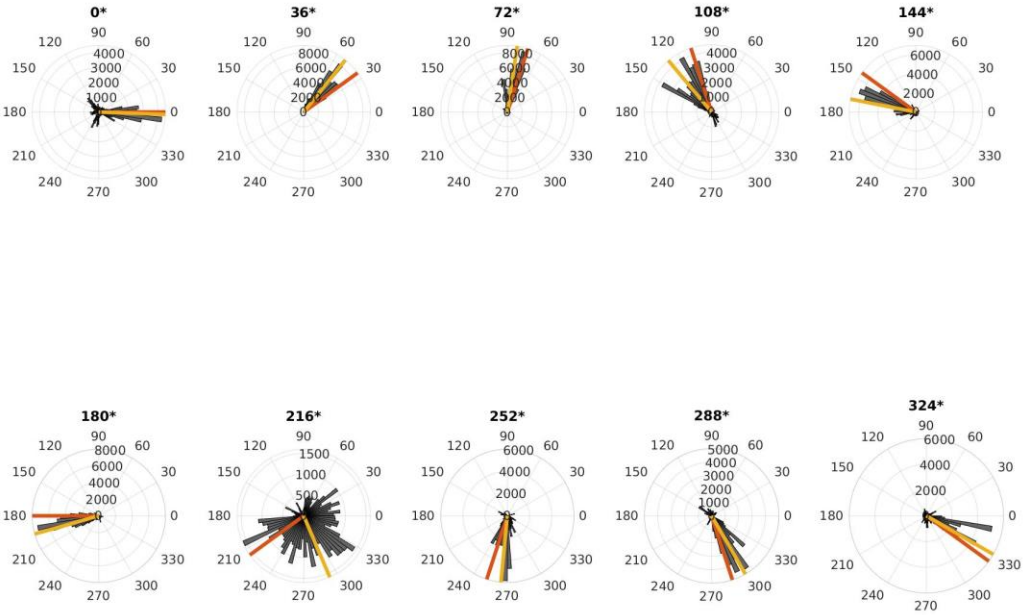

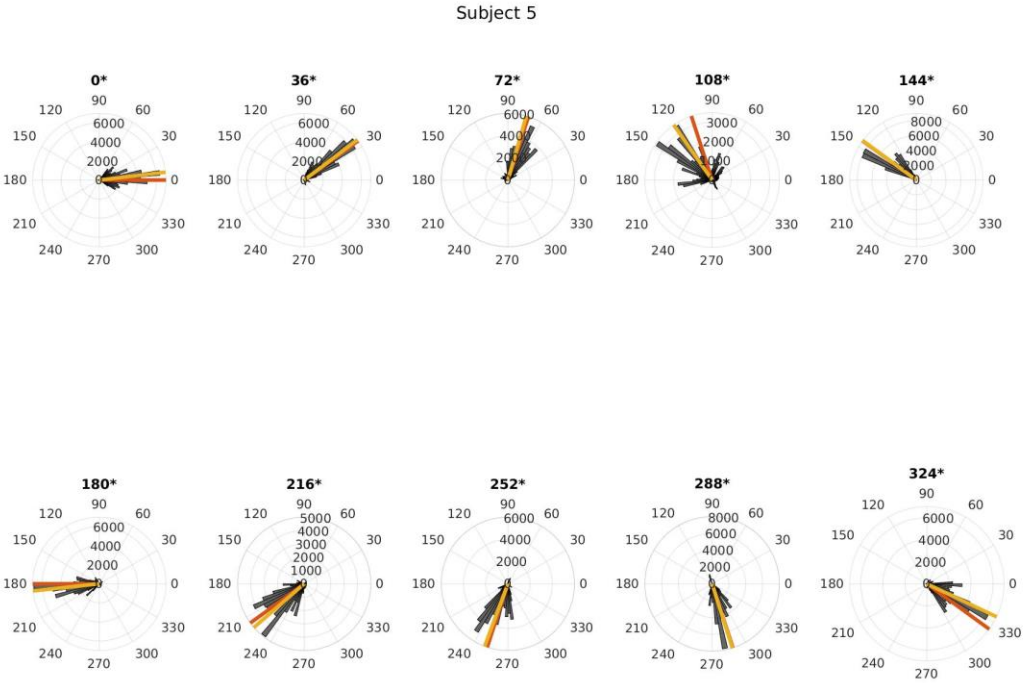
Subject 5
0*
36*
72*
108*
144*
180*
216*
252*
288*
324*

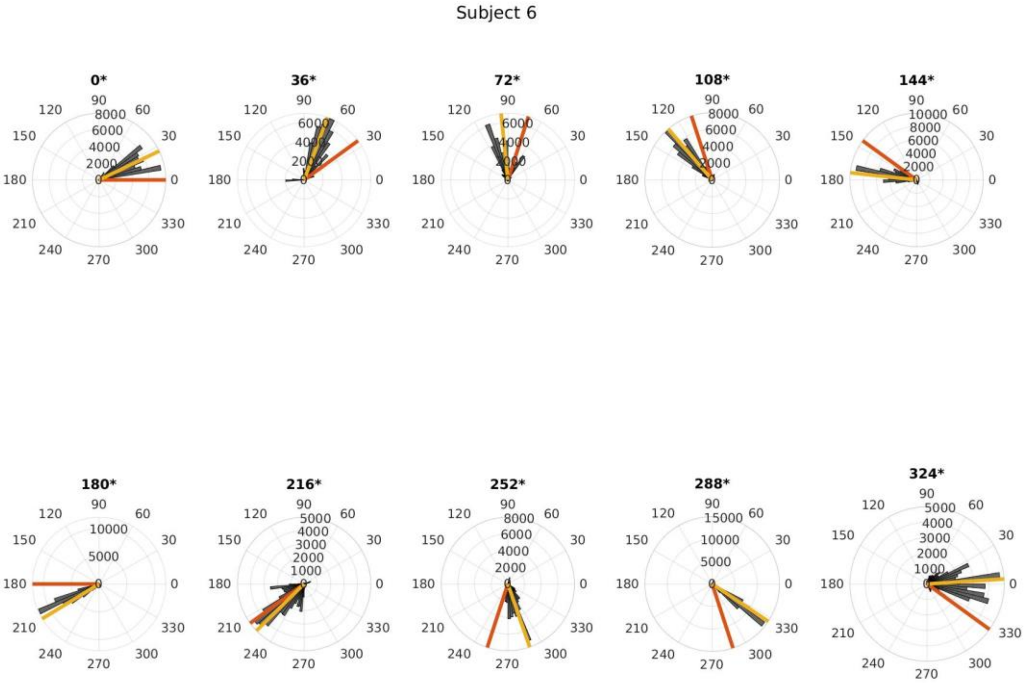
Subject 6
0*
36*
72*
108*
144*
180*
216*
252*
288*
324*

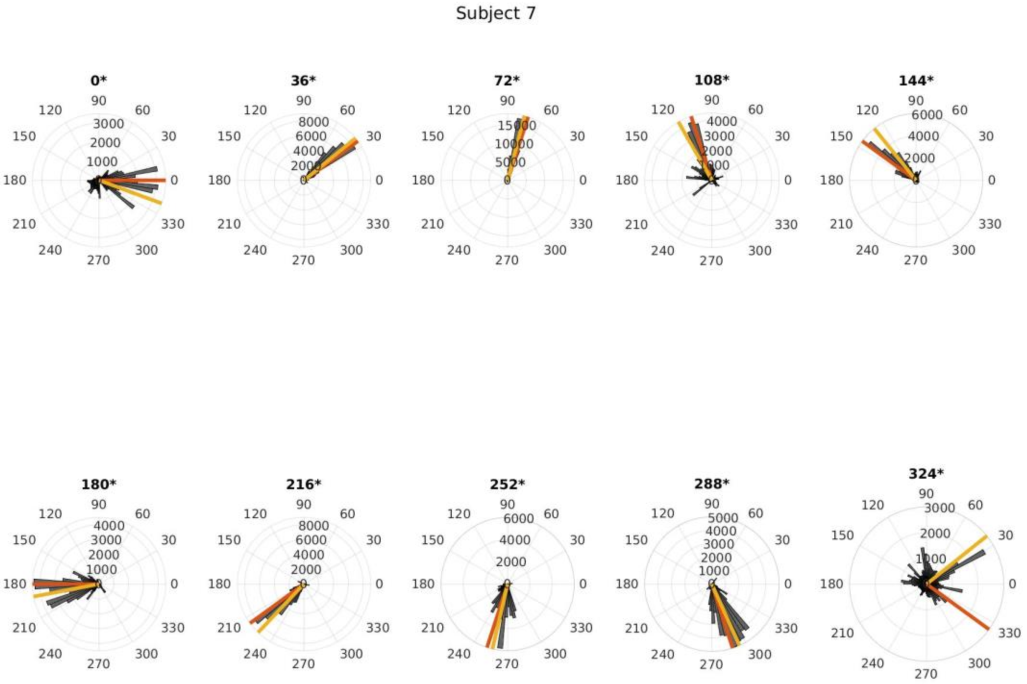
Subject 7
0*
36*
72*
108*
144*
180*
216*
252*
288*
324*

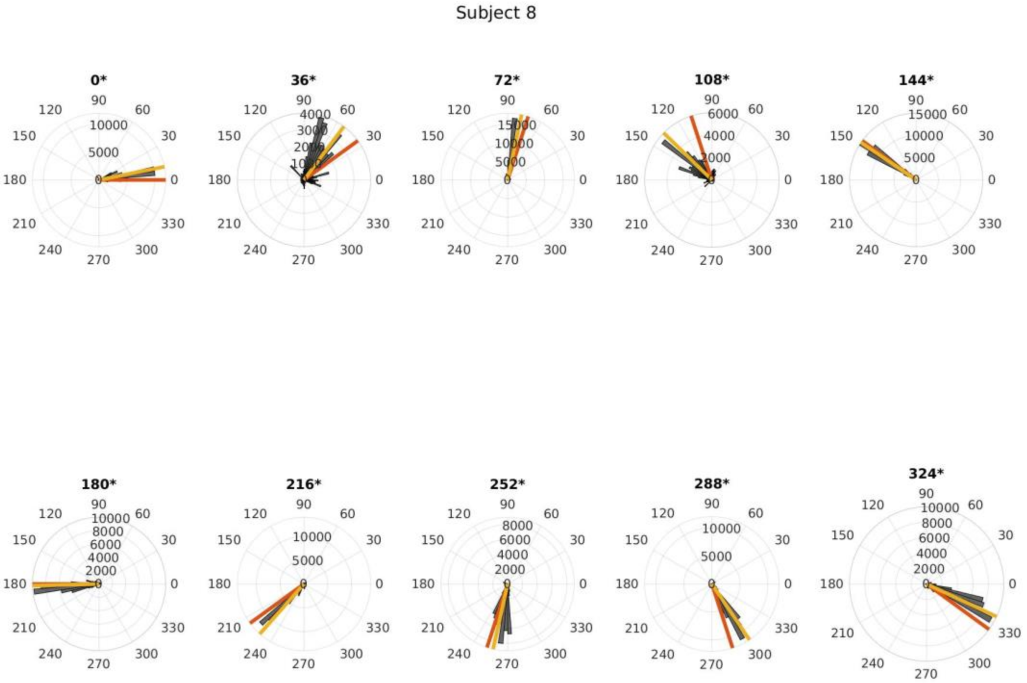
Subject 8
0*
36*
72*
108*
144*
180*
216*
252*
288*
324*

Subject 9

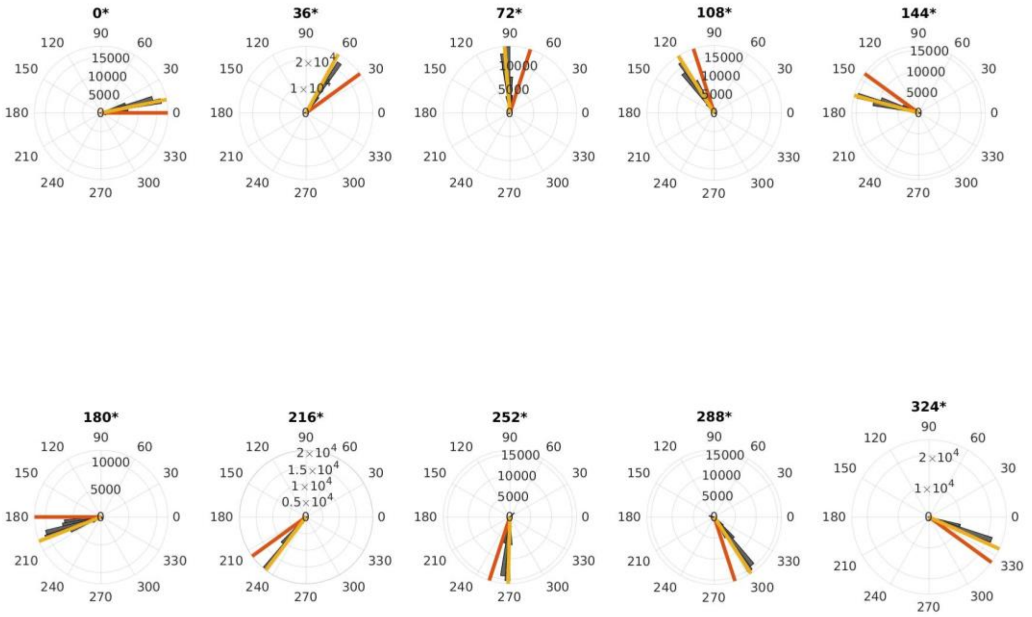


Subject 10

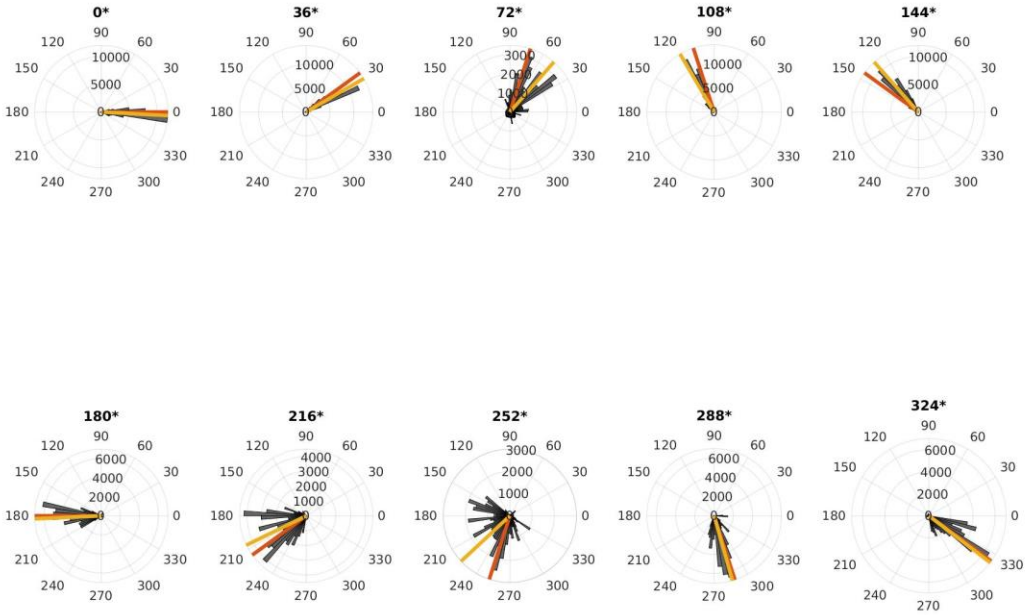

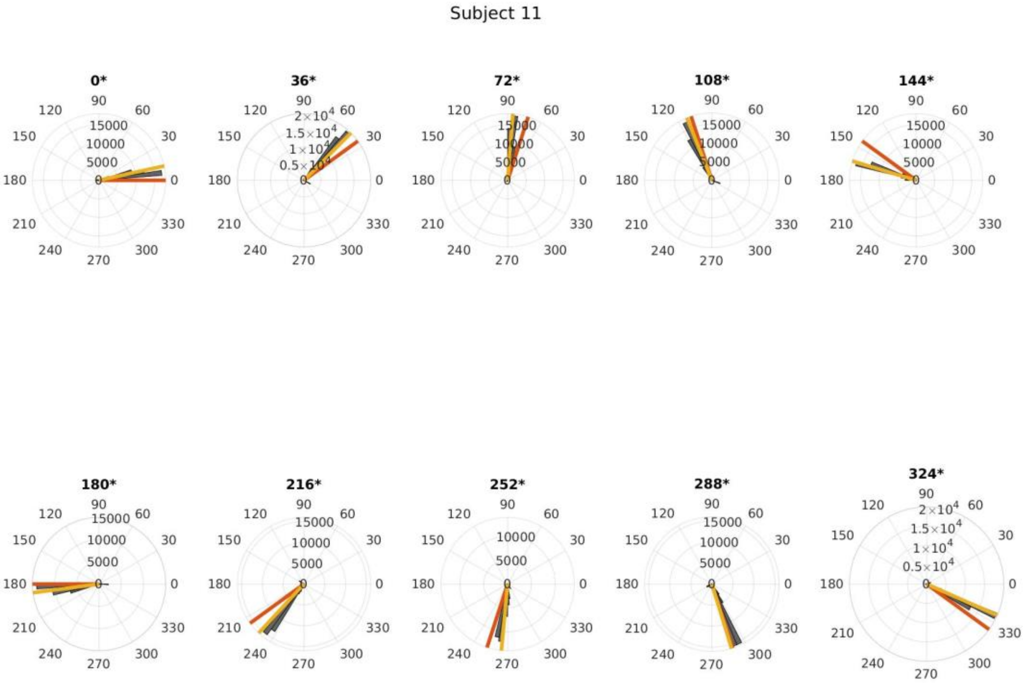
Subject 11
0*
36*
72*
108*
144*
180*
216*
252*
288*
324*

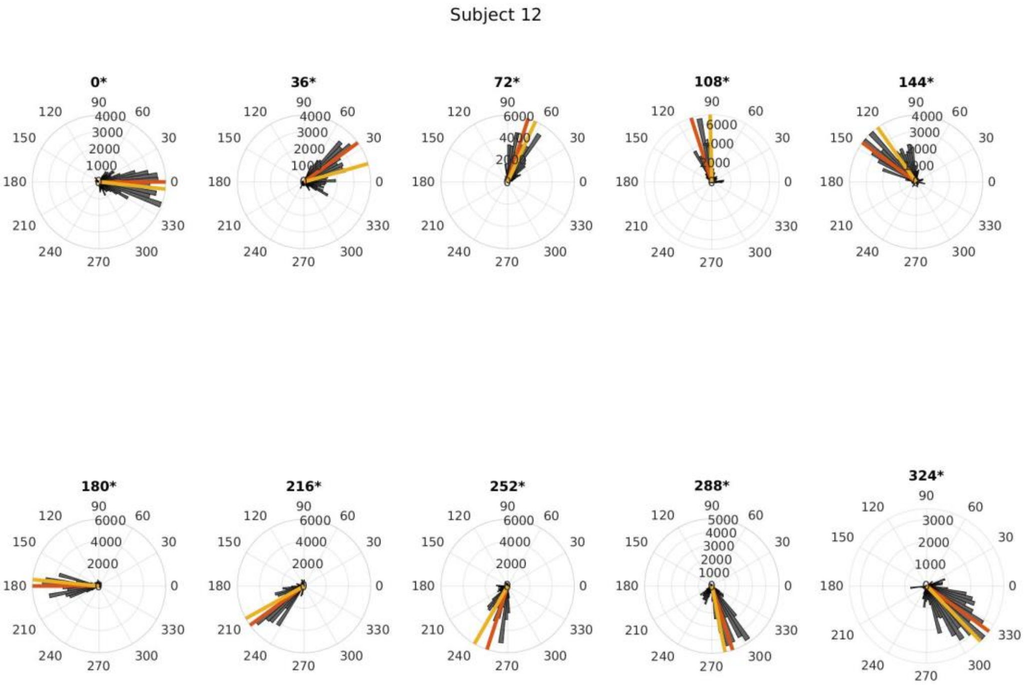
Subject 12
0*
36*
72*
108*
144*
180*
216*
252*
288*
324*

Subject 13

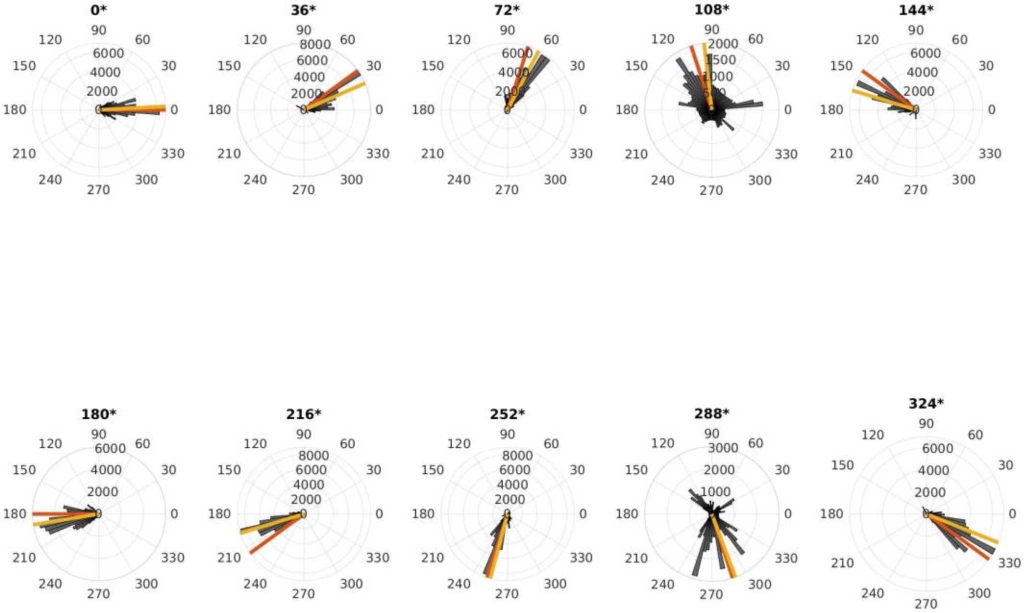


Subject 14

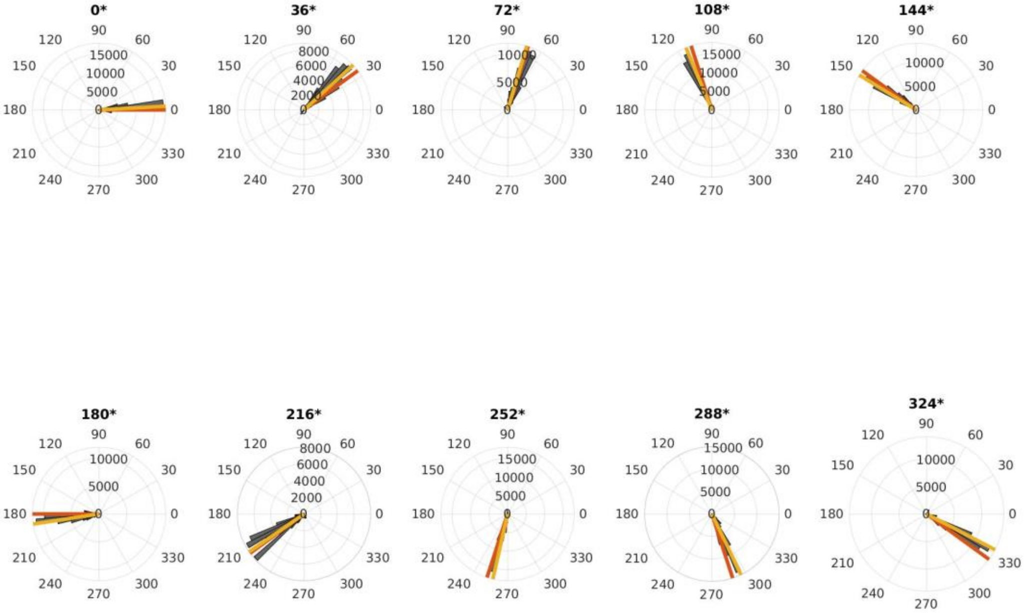

Subject 15

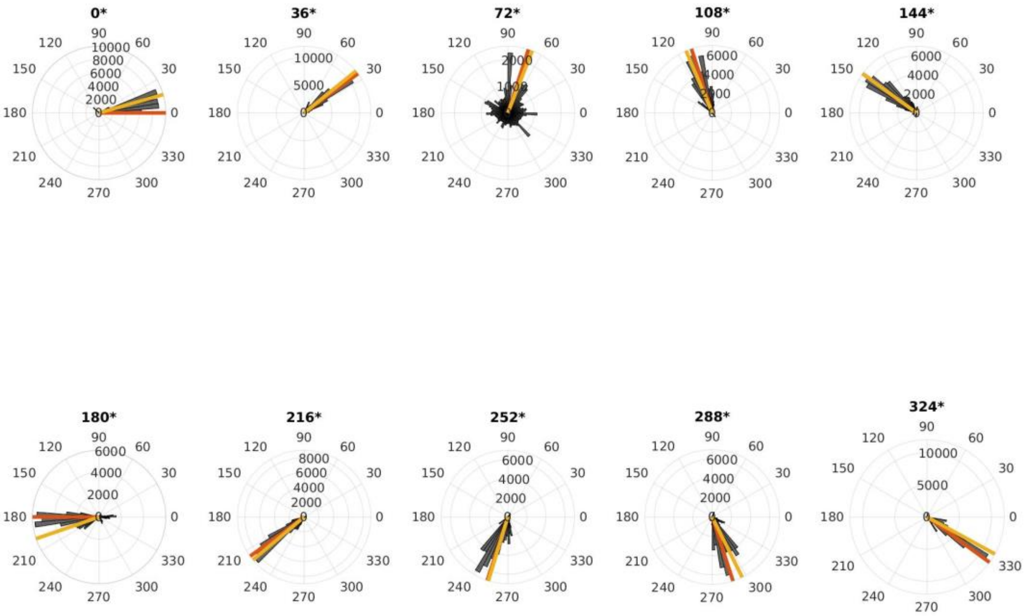


Subject 16

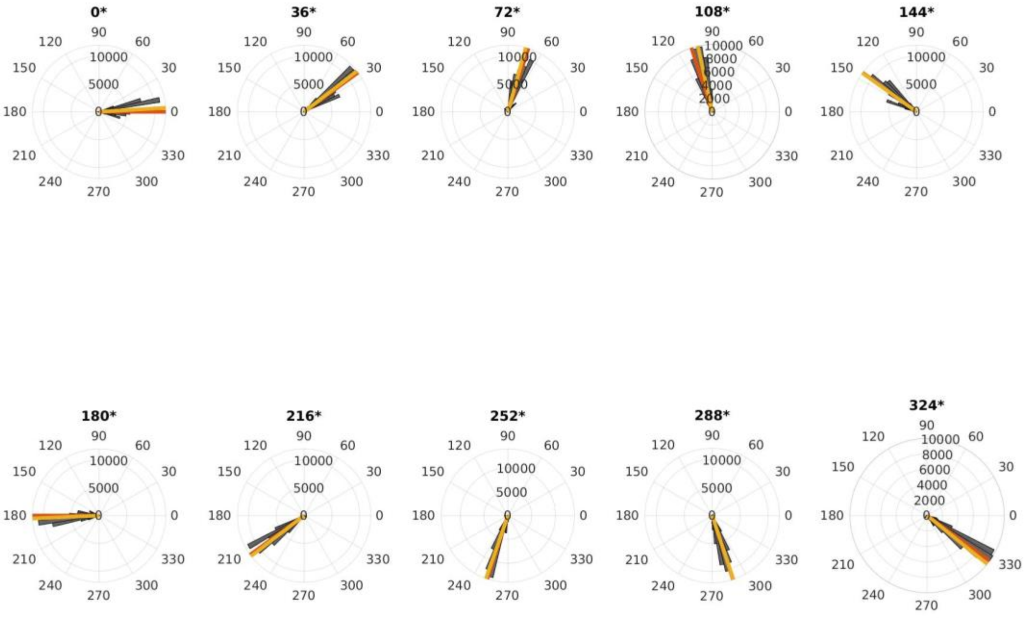

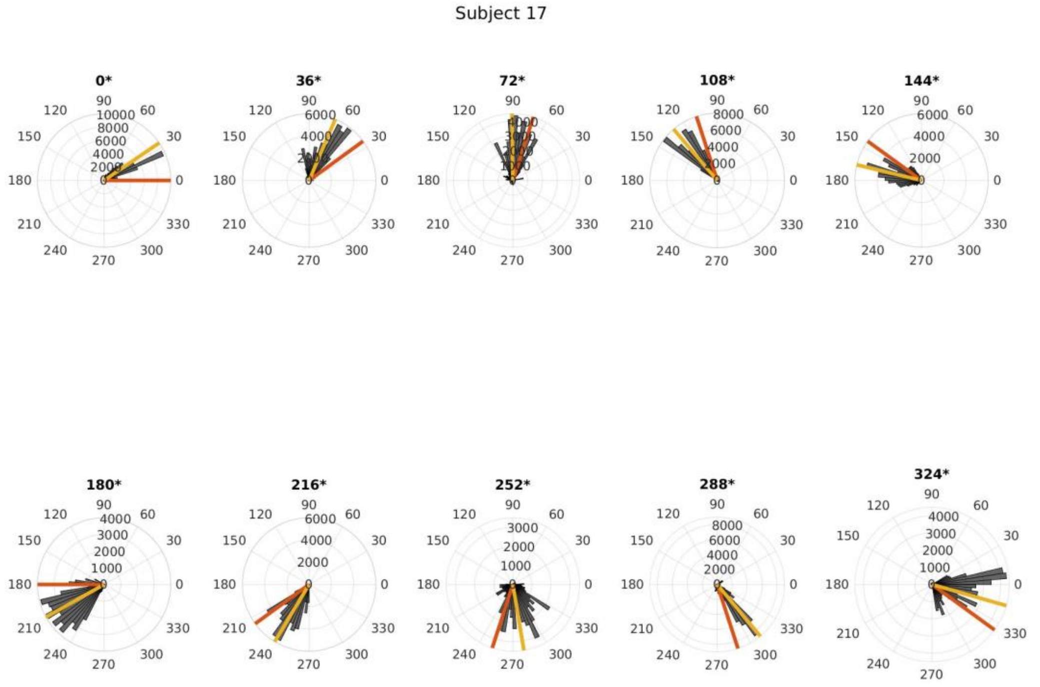
Subject 17
0*
36*
72*
108*
144*
180*
216*
252*
288*
324*

**Supplementary figures: Phase adequacy of phase-locked transcranial alternating current stimulation of the cerebellum.**

Figures on the next 7 pages show the adequacy of phase-locked stimulation for cerebellum stimulation. Figure layout is identical to figure 2 of the main manuscript.

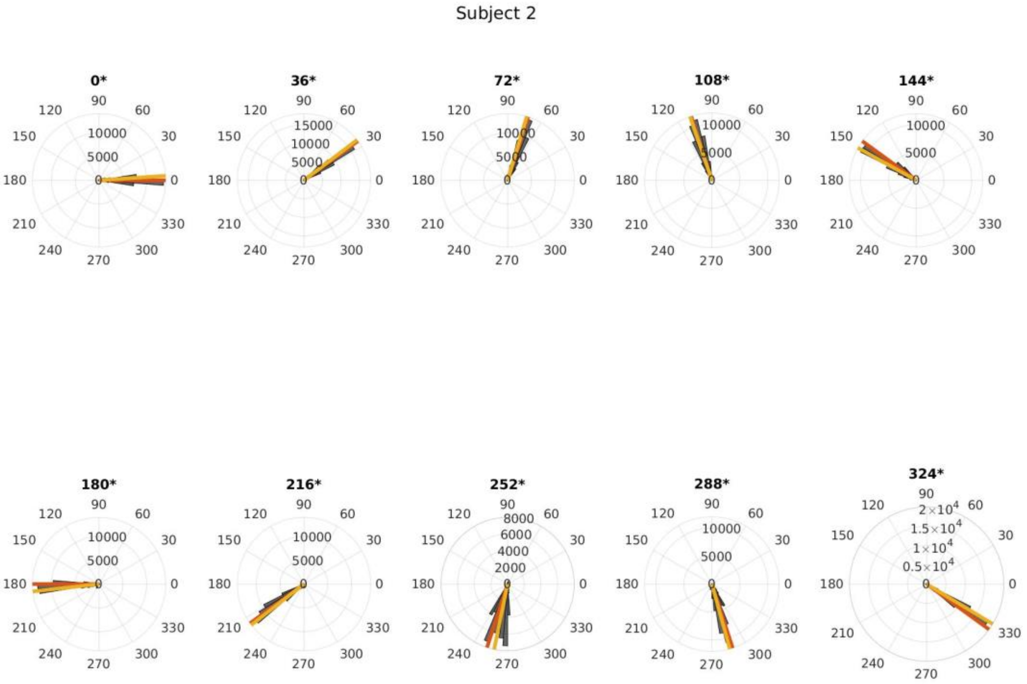
Subject 2
0*
36*
72*
108*
144*
180*
216*
252*
288*
324*

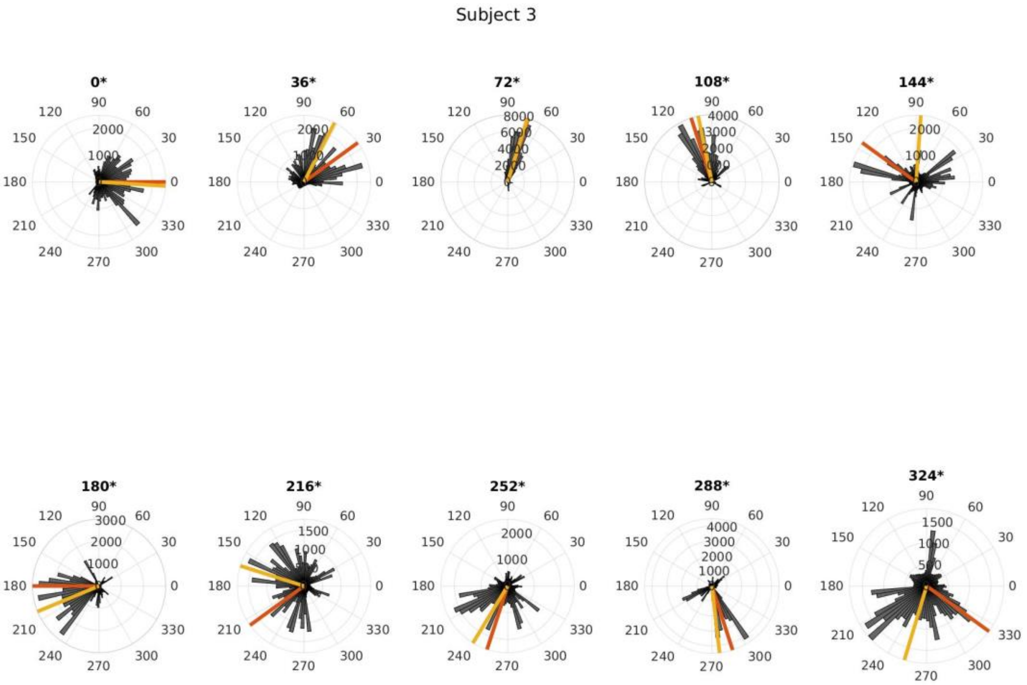
Subject 3
0*
36*
72*
108*
144*
180*
216*
252*
288*
324*

Subject 4

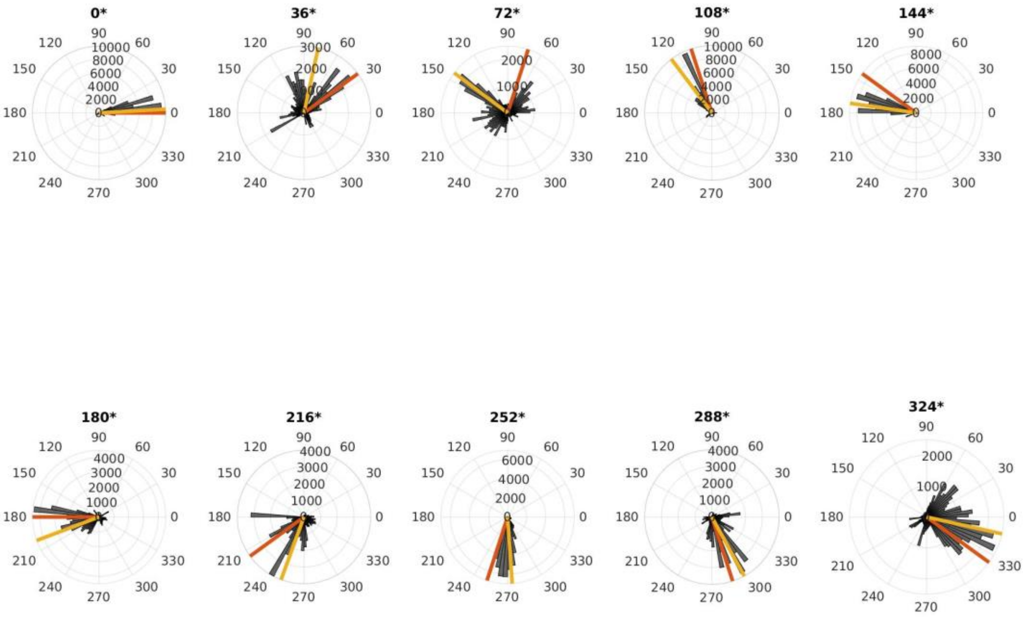


Subject 5

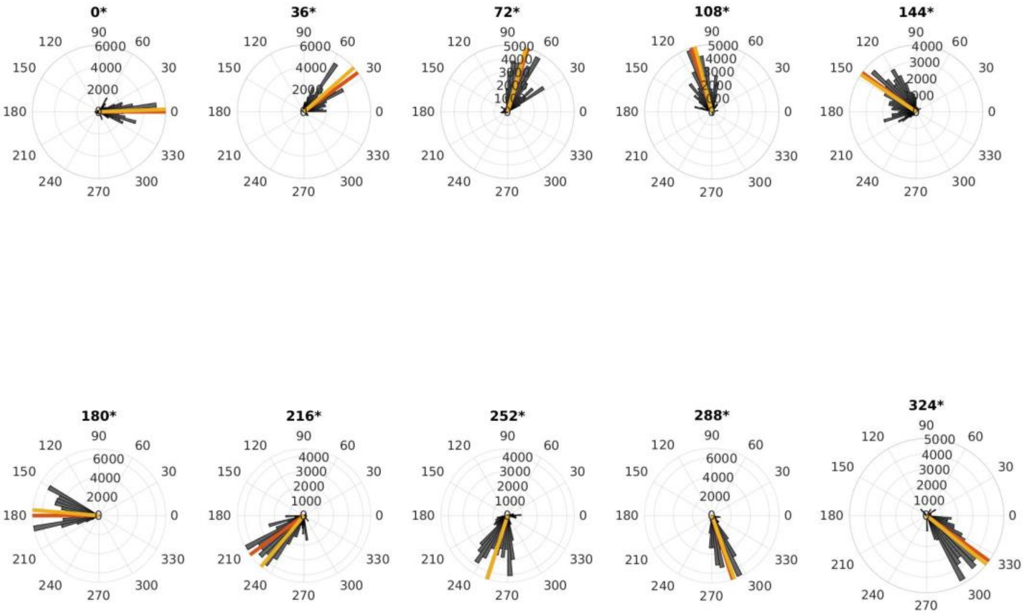

Subject 6

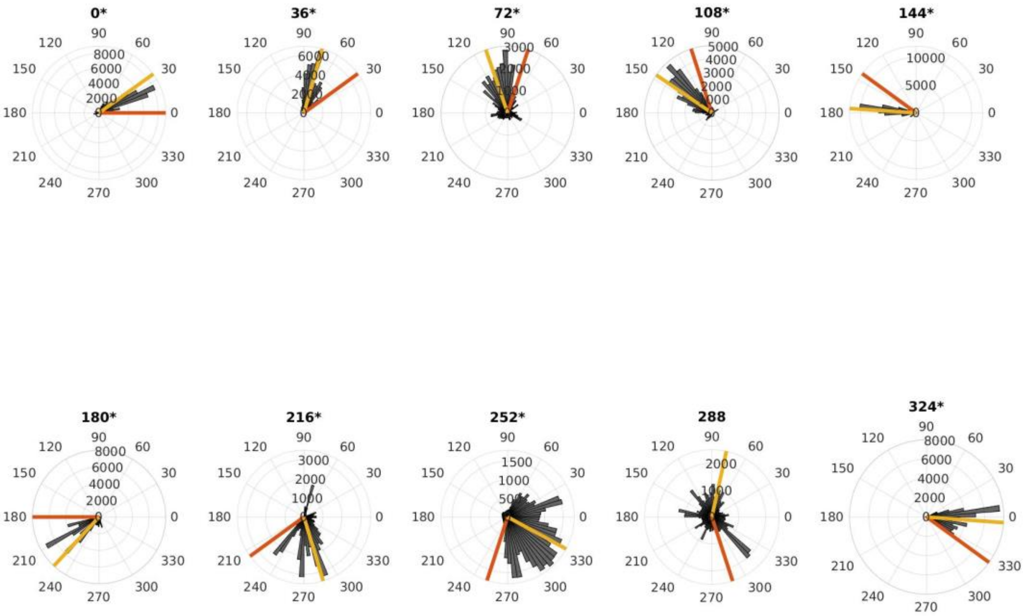


Subject 7

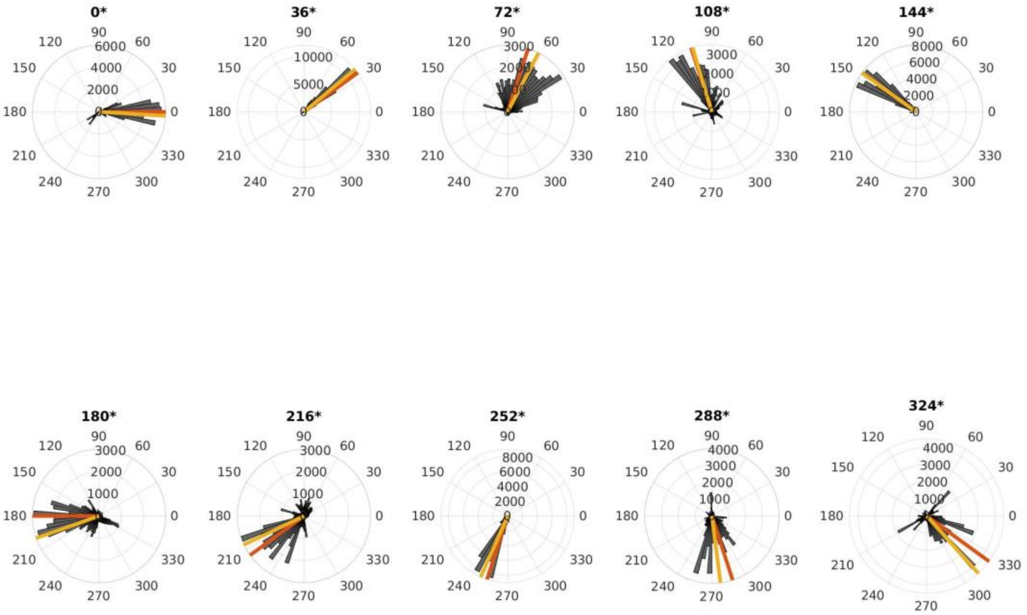

Subject 8

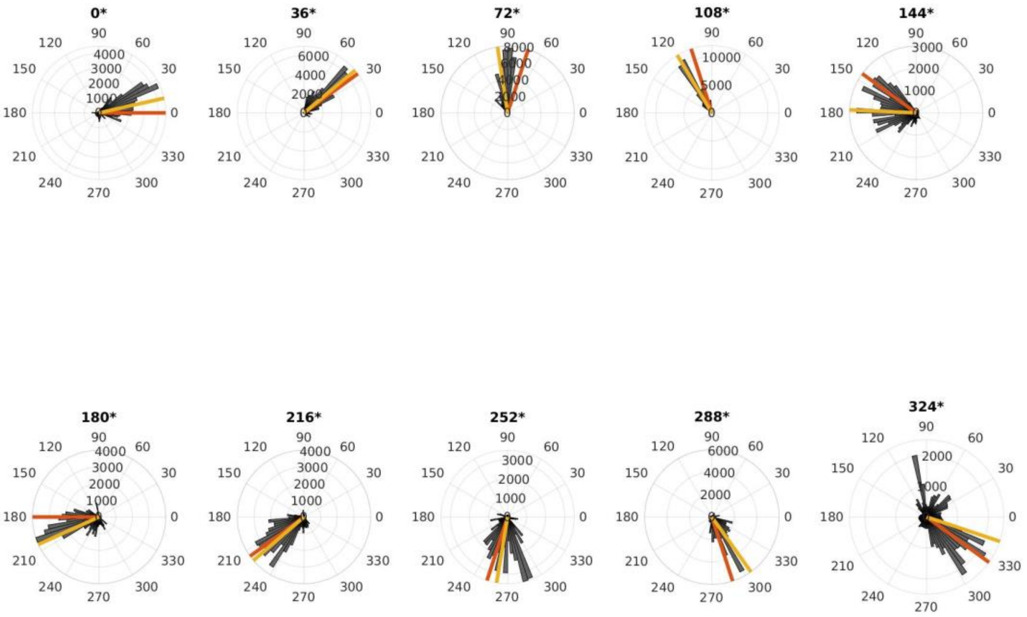


Subject 9

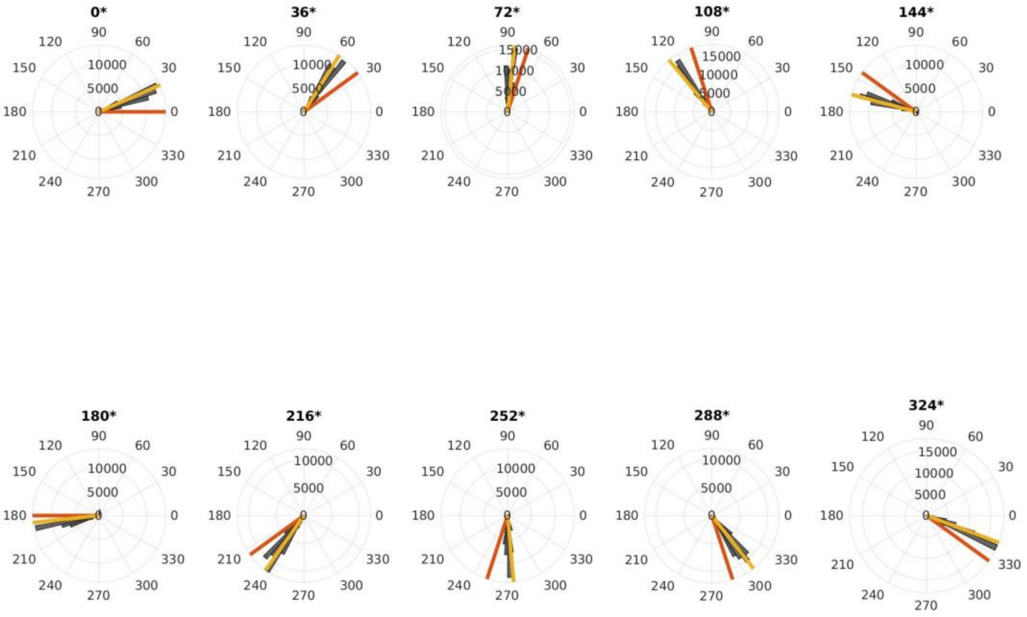

Subject 11

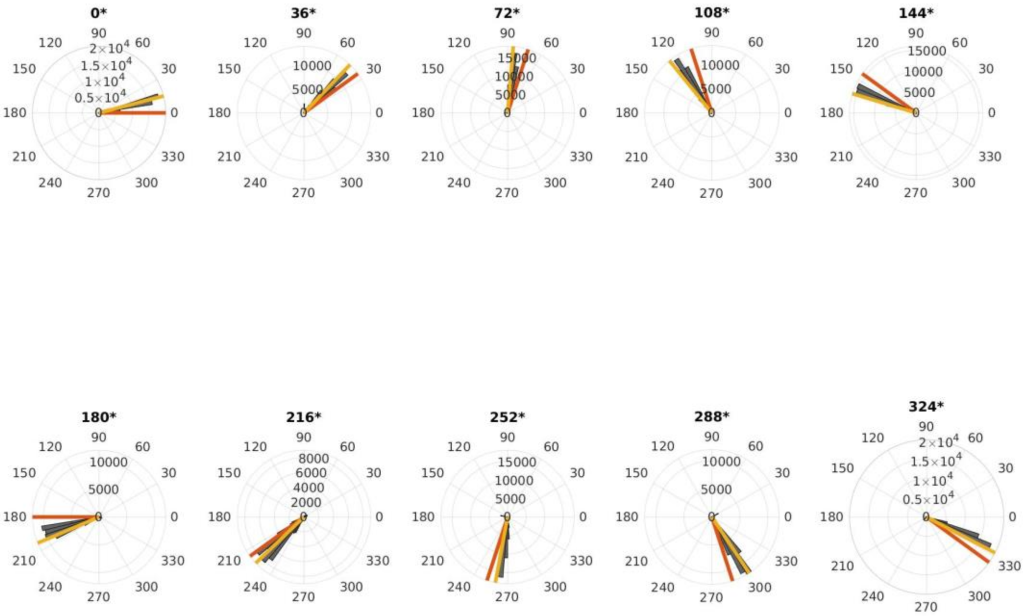


Subject 13

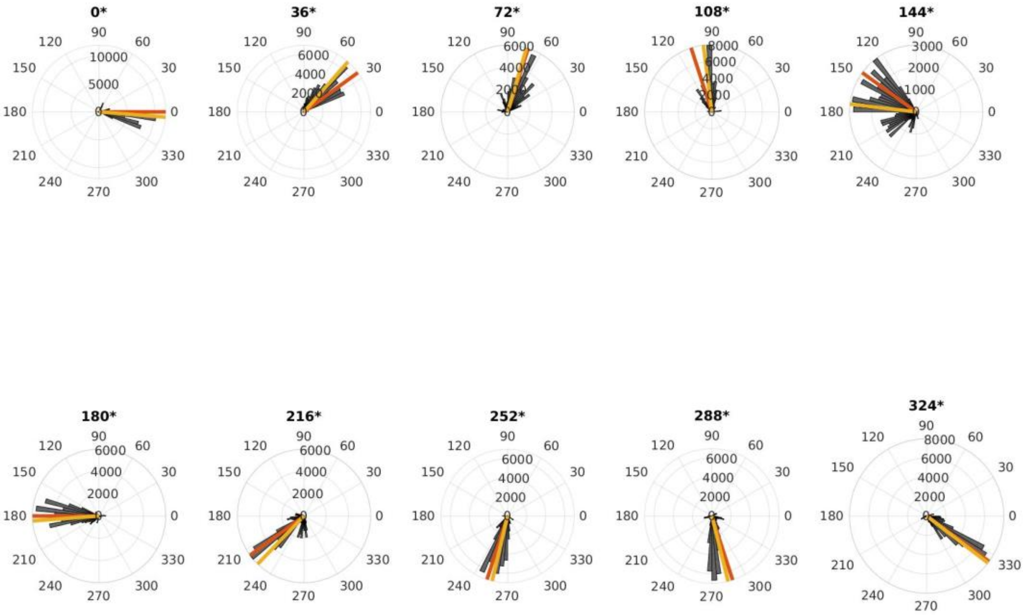

Subject 14

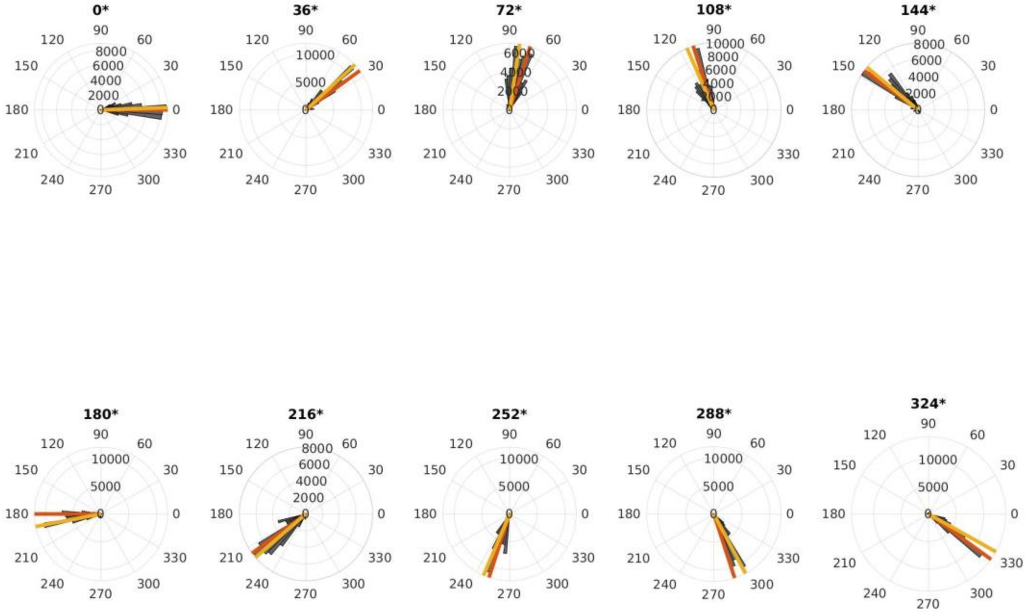


Subject 15

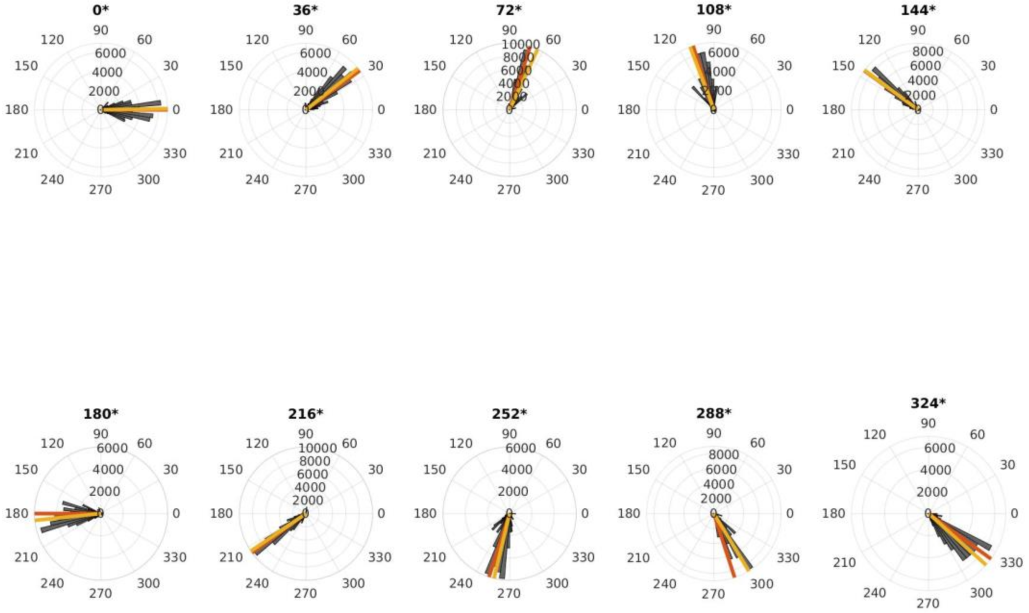

Subject 16

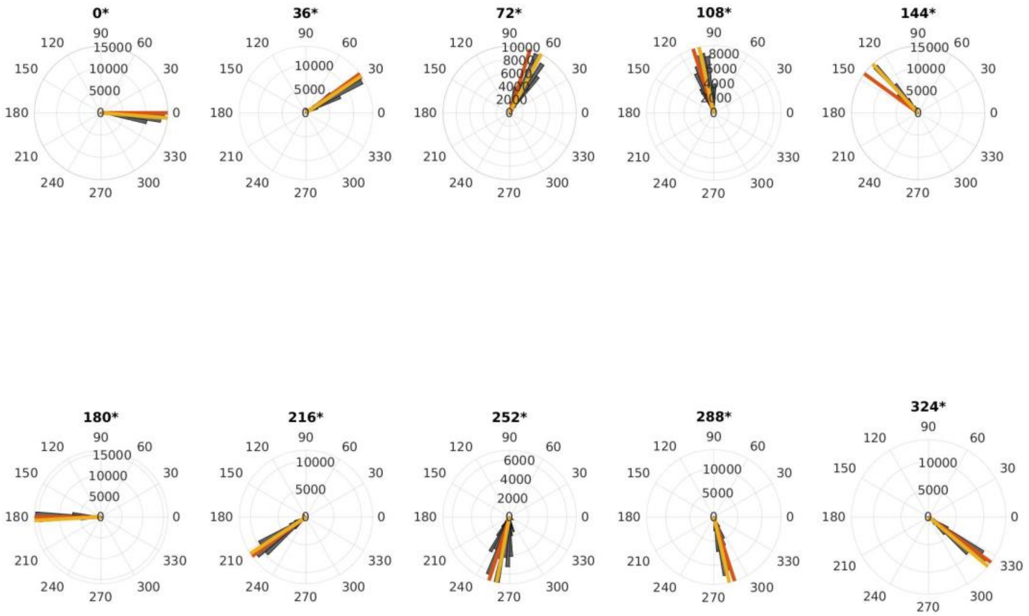


Subject 17

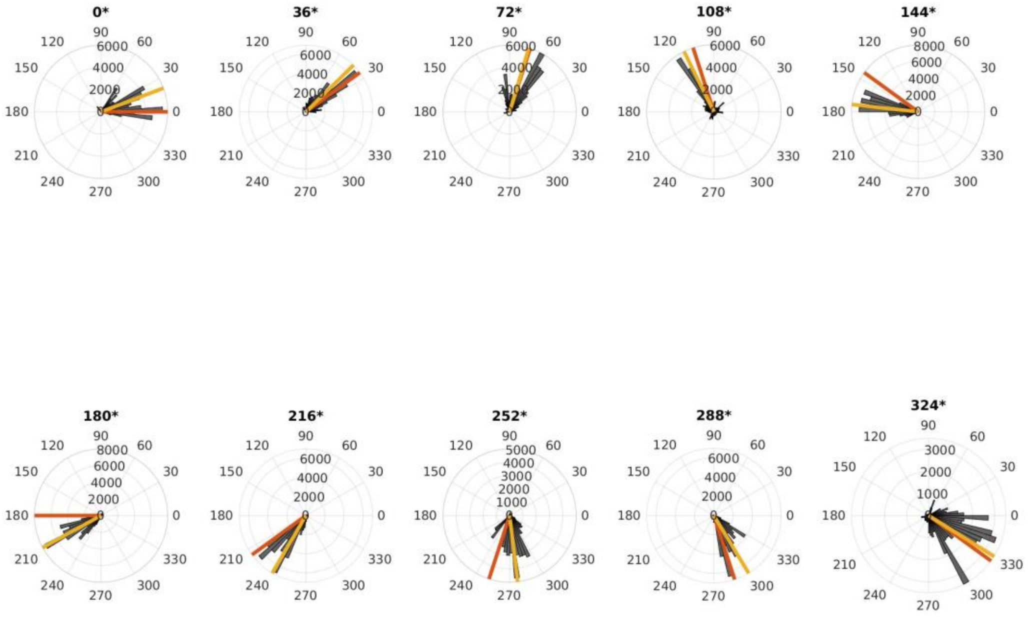
